# Prevalence of high-risk human papillomavirus genotypes and associated HIV co-infection in Cameroon: a systematic review and meta-analysis

**DOI:** 10.64898/2026.03.10.26348064

**Authors:** Fabrice Zobel Lekeumo Cheuyem, Chabeja Achangwa, Lionel Berthold Keubou Boukeng, Rick Tchamani, Alapa Junior Nkwate Chefor, Jessy Goupeyou-Youmsi, Edmond Martial Lemaire Bodo

**Author notes:** Corresponding author’s address: Fabrice Zobel Lekeumo Cheuyem.

## Abstract

**Background:** High-risk human papillomavirus (HR-HPV) infection is the necessary cause of cervical cancer, a leading cause of cancer mortality among women in sub-Saharan Africa. In Cameroon, there is a gap in synthetized evidence on HR-HPV epidemiology, limiting data-driven prevention strategies. This study provides the first comprehensive national synthesis of HR-HPV prevalence, genotype distribution, and HIV co-infection among HPV-infected individuals.

**Methods:** A systematic review and meta-analysis were conducted following PRISMA guidelines and registered in PROSPERO (CRD420261282094). We systematically searched PubMed, Scopus, Web of Science, Embase, Cochrane Library, AJOL and local online publishers. Studies reporting the prevalence of HR-HPV among sexually active individuals were included. Pooled prevalence estimates were calculated using random-effects meta-analysis. Heterogeneity was assessed with *I²* statistics and explored through subgroup analyses. The methodological quality of included studies was evaluated using the Joanna Briggs Institute (JBI) critical appraisal tools.

**Results:** A total of 33 studies including 18,798 participants were included in this analysis. The overall pooled prevalence of HR-HPV was 28.95% (95% CI: 19.74–40.30; 33 studies; n = 18,798; *I*^2^ = 98.7%), with a notable decline from 53.34% before 2014 to 21.43% in 2021–2023. Prevalence varied substantially across populations, being highest among women with precancerous or cancerous lesions (90.69%; 95% CI: 78.48–96.30) and female sex workers (62.10%; 95% CI: 58.08–66.00) compared to women from the general population (21.09%; 95% CI: 15.16–28.55). Among HPV-positive women, HIV co-infection prevalence was 26.17% (95% CI: 13.48–44.65, 19 studies; n = 3,589; *I*^2^ = 97.9%), with higher rates in hospital-based studies (34.57%) compared to community-based studies (9.18%). Predominant HR-HPV genotypes included HPV16 (28.7%), HPV52 (23.6%), HPV6 6 (22.9%), HPV33 (22.8%), and HPV18 (20.2%). The pooled prevalence of abnormal cervical lesions among HPV-positive women was 35.15% (95% CI: 20.21–53.70; 12 studies; n = 2,186; *I*^2^ = 95.2%), comprising low-grade lesions (34.4%) and high-grade lesions (19.1%).

**Conclusions:** High-risk human papillomavirus infection remains highly prevalent in Cameroon despite encouraging temporal declines. The substantial burden of HIV co-infection and circulation of multiple oncogenic genotypes underscore the need for integrated HPV–HIV prevention strategies, expanded screening, and consideration of broader-coverage vaccines to accelerate progress toward cervical cancer elimination targets.

## 1. Introduction

Infection with high-risk human papillomaviruses (HR-HPV) is the most widespread sexually transmitted viral infection in the world and the primary etiological cause of cervical cancer [1]. Approximately 660,000 new cases and 350,000 deaths from cervical cancer have been recorded worldwide in 2024, making this pathology the fourth leading cause of cancer in women and the second in those aged 15 to 44 years [2]. Sub-Saharan Africa bears a high burden, with incidence and mortality rates among the highest in the world, reaching 40 cases and 28 deaths per 100,000 women per year, respectively [3]. This situation is primarily due to a high prevalence of HR-HPV infections and limited access to screening and treatment services for precancerous lesions, as well as still insufficient vaccination coverage in the region [2]. Persistent infection with HR-HPV, particularly genotypes 16, 18, 31, 33, 45, 52, and 58, is recognized as responsible for more than 99% of cervical cancer cases, thus justifying the importance of its epidemiological surveillance [4].

In Cameroon, studies conducted revealed HR-HPV prevalence rates among populations ranging from 4.2% to 52.5% among women in the general population [5]. These rates were notably higher among women living with HIV (38.5% to 52.9%) and sex workers (62.1%) [6, 7]. The distribution of genotypes reveals a predominance of types 16, 18, 52, and 58, with geographical variations and some genotypes not covered by current vaccines [5]. The coexistence of HIV, with a national prevalence of 3.3% among women, complicates the epidemiological landscape by increasing the risk of viral persistence and progression to precancerous lesions [6, 8].

The literature presents significant disparities in the estimates of HR-HPV prevalence, likely related to methodological differences between studies. Moreover, the available data on HR-HPV infections has not yet been aggregated, limiting therefore the national estimation of the true prevalence. This situation limits the understanding of the epidemiology of HR-HPV and does not allow identification of disparities to guide targeted public health interventions. The availability of reliable and national estimates is essential to inform public health policies, particularly in the context of integrating the HPV vaccine into the Expanded Program on Immunization schedule since 2020. Thus, a better understanding of the prevalence of HR-HPV, circulating genotypes, co-infection with HIV, and associated cervical lesions would allow for the adaptation of interventions to the most vulnerable populations, provide baseline data to assess the future impact of prevention programs, and optimize resource allocation. The present study aimed to estimate the prevalences of HR-HPV genotypes and HIV among HPV-infected individuals in Cameroon.

## 2. Methods

### 2.1. Study design

This review followed Preferred Reporting Items for Systematic Reviews and Meta-Analyses (PRISMA) reporting standards and was registered in the International Prospective Register of Systematic Reviews under ID CRD420261282094.

### 2.2. Eligibility criteria

#### Inclusion criteria

Studies were eligible for inclusion if they reported on the prevalence of HPV among sexually active individuals in Cameroon. Both observational and interventional study designs were considered. We included articles published in English and French, with no restrictions applied to the date of study.

#### Exclusion criteria

Studies with duplicated information, with research topics not related to the objective of our study, case reports, posters, conference papers, reviews, editorials, commentaries, and animal-based studies were not considered for this study. In addition, studies lacking sufficient data to calculate prevalence estimates were excluded. Multi-country studies without extractable Cameroon-specific data were also excluded.

### 2.3. Search Strategy

Relevant studies were identified through a systematic search of global and regional electronic databases. Global databases included PubMed, Scopus, Web of Science, Embase, and the Cochrane Library, while regional context was ensured by searching AJOL and the Health Sciences and Disease journal. To minimize publication bias, we included grey literature including preprints by conducting manual search on in Google Scholar. This was finalized by the screening of bibliographies from identified eligible articles.

To identify relevant literature effectively, we employed a search strategy combining Medical Subject Headings (MeSH) terms and Boolean operators. The specific search string used in PubMed was as follows: (“human papillomavirus”[tiab] OR HPV[tiab] OR “cervical cancer”[tiab] OR “uterine cervical neoplasms”[Mesh]) AND (Cameroon[tiab] OR “Cameroon”[Mesh]) (**Additional file 1, Supplementary Table 1**). Screening was conducted by two investigators (FZLC and RT), and any disagreements were resolved through discussion to reach a consensus. The literature search was completed on January 27, 2025.

### 2.4. Data extraction

We extracted key data from each study into a Microsoft Excel spreadsheet. For each study, we recorded the following information: the first author’s last name, the year of study completion, the region of Cameroon where the study was conducted, the type of participants, the sampling method, the diagnostic tool used, the sample size, the number of eligible participants screened for HPV, the number of HR HPV cases detected, the number of HPV-positive cases screened for HIV, the number of HIV-positive cases diagnosed. For each eligible studies, we also extracted in a separated spreadsheet the number of HR-HPV genotype identified, cytologic or histologic uterine cervical lesions observed among HPV-positive women and the total number of HPV-positive women. This step was performed by one investigator (FZLC) and cross-checked by two other research team members (CA and LBKB). A discussion was engaged between study investigators to solve any observed discrepancies.

### 2.5. Study outcomes and operational definition

Our study primary outcome was the High-risk prevalence of HPV in Cameroon. The secondary outcomes included the prevalence of HIV and abnormal cervical lesions among HPV-positive women. An HPV-positive case was defined as any participant with a valid sample collected either by a healthcare provider or through self-sampling that tested positive via genotyping or an equivalent validated laboratory test. HR-HPV-positive cases referred to any HPV-positive individuals whose genotyping test identified at least one of the following oncogenic serotypes (31, 33, 35, 39, 51, 52, 56, 58, 59, 66, and 68) [5, 7, 9, 10]. A case was defined as laboratory-confirmed HIV if at least one specimen showed a positive result in an HIV antigen-specific assay or HIV-specific real-time PCR [5, 7, 10, 11].

### 2.6. Risk of bias assessment

Methodological quality evaluation was carried out by two independent reviewers (FZLC and CA) using the Joanna Briggs Institute (JBI) critical appraisal tools, adapted for each specific study design. Any discrepancies were resolved via consensus or by consulting a third reviewer (EMLB). For cross-sectional studies, the assessment focused on the clarity of inclusion criteria, the detail of subject and setting descriptions, the validity of exposure measurements, and the objectivity of outcome assessments. We also evaluated the identification of confounding factors, the strategies used to mitigate them, and the rigor of the statistical analyses. Each criterion was scored as 1 (Yes) or 0 (No/Unclear). The overall risk of bias was categorized based on the percentage of ‘Yes’ scores: low (>50%), moderate (26–50%), or high (≤25%).

### 2.7. Data analysis

We computed each prevalence estimates along with 95% confidence intervals (CI). To account for expected high heterogeneity, we used a random-effects meta-analysis (DerSimonian-Laird method) to generate pooled prevalence estimates. We will assess statistical heterogeneity using the *I²* statistic and Cochran’s Q test. To assess potential source of heterogeneity, subgroup analyses was conducted on study period, geographic region, population type, study design, and sampling method used. The Generalized Linear Mixed Models (GLMM), coupled with the Probit-Logit Transformation (PLOGIT) were two combined-methods used to generate study pooled estimates. They were choose because of their effectiveness in handling meta-analyses of binary data [12]. Statistical significance was set at a *p*-value of <0.05. All analyses were conducted using the ‘meta’ package in R Statistics version 4.5.2 [13].

### 2.8. Publication bias and sensitivity analysis

Publication bias was evaluated through visual inspection of funnel plots and confirmed with formal statistical tests, including Egger’s and Begg’s tests [14, 15]. A *p*-value below 0.05 was used to indicate significant bias. Furthermore, we conducted a sensitivity analysis using the ‘leave-one-out’ approach—iteratively excluding one study at a time—to evaluate the stability and robustness of our pooled estimates.

## 3. Results

The systematic search identified 741 records (733 from databases and 8 from other sources). After removing 43 duplicates, we screened 698 records by title and abstract and then assessed 57 by full-text. Of these, 33 primary research articles met the eligibility criteria and were included in the systematic review and meta-analysis (**Fig. 1**).

**Fig. 1.**
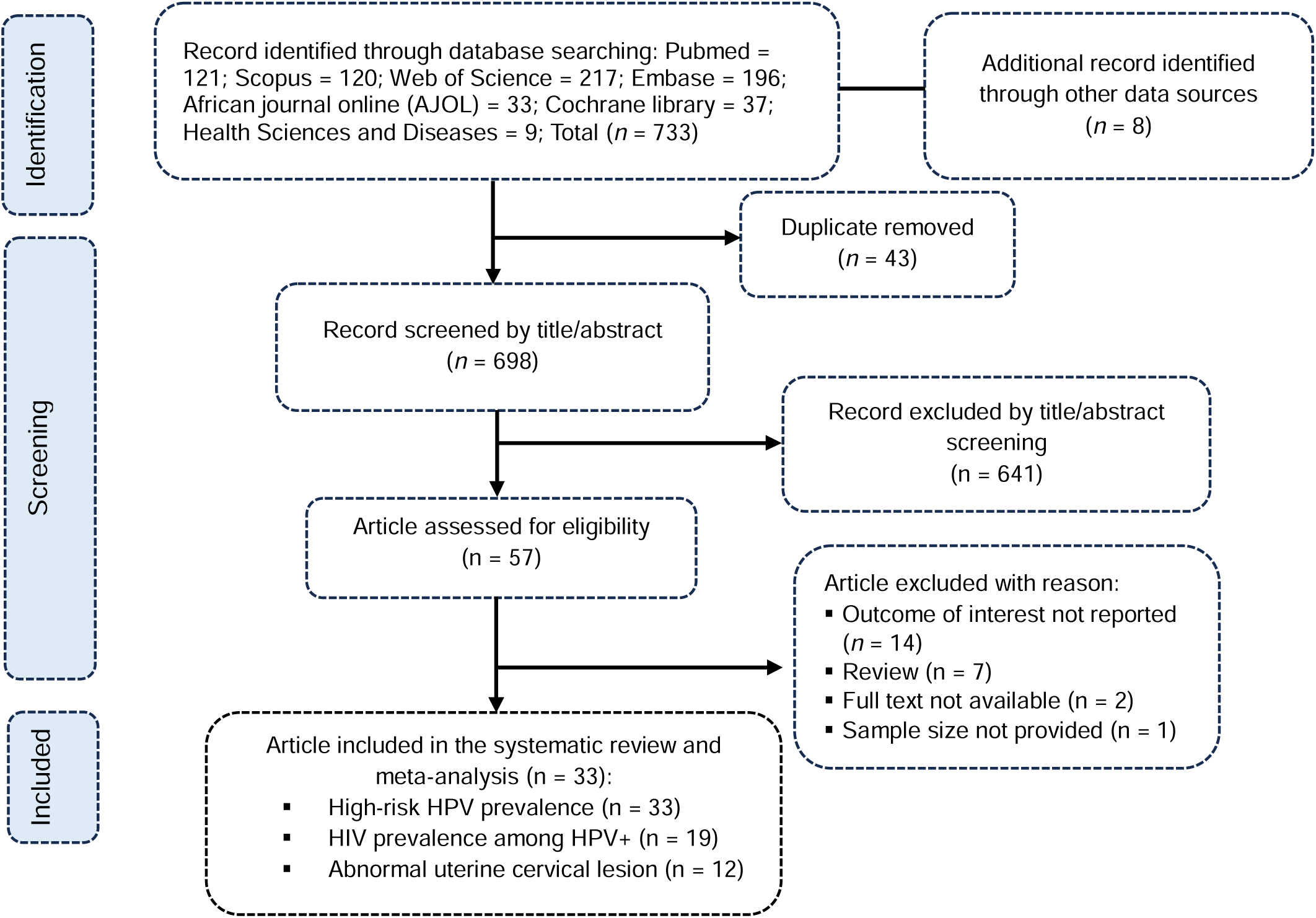
PRISMA diagram flow from study identification to inclusion in the meta-analysis (*HPV: Human papillomavirus*)

### 3.1. Studies selection

### 3.2. Study characteristics

A total of 33 studies measured high-risk HPV prevalence in Cameroon and 19 assessed HIV prevalence among HPV-positive cases. Most of included studies were a cross-sectional (n = 31; 93.9%). Nearly half of all studies were conducted in the Centre region (n = 16; 48.5%), followed by the Littoral (n = 9; 27.3%) and the West regions (n = 8; 24.2%); other studied regions included the South-West (n = 5; 15.2%) and the North-West regions (n = 3; 9.1%). Nearly three-quarters of studies (n = 24; 72.7%) were conducted exclusively in hospital settings and 24.2% (n = 8) in community. Three-quarters of studies (n = 25; 75.8%) targeted the general female population including sexually active pregnant and non-pregnant women. A small proportion focused on high-risk subgroups, including women with cervical lesions (n = 3; 9.1%), people living with HIV (n = 3; 9.1%). Most studies used a non-probabilistic sampling method to include participants (n = 31; 93.9%) (**Table 1**).

**Table 1.** Characteristics of studies included.

| Author | Year<sup>1</sup> | Design | Region | Setting | Sampling method | Type of participant | Age (years ) | Median IQR or Mean SD | HPV diagnostic tool | Risk of bias | High-risk HPV<sup>2</sup> HIV<sup>3</sup> (Event/n) |
| --- | --- | --- | --- | --- | --- | --- | --- | --- | --- | --- | --- |
| Desruisseau,2006 [10] | 2006 | Cross-sectional | Centre and South-West | Hospital | Non-probalistic | Non-pregnant women | 18+ | NR | PCR-based reverse-line strip test for multiple HPV types | Low | 32/61 = 52.46<br>26/41 = 63.41 |
| Sando,2009 [16] | 2009 | Cross-sectional | Centre | Hospital | Non-probalistic | Women with pre/cancerous genital lesion | 28-55 | NR | PCR-based reverse-line strip test for multiple HPV types | Low | 28/37 = 75.68<br>NR |
| Teubeu,2010 [17] | 2010 | Cross-sectional | West | Hospital | Non-probalistic | Women | 30-75 | NR | Abbott RealTime High-Risk HPV assay | Low | 26/157 = 16.56<br>NR |
| Pirek,2010 [18] | 2010 | Cross-sectional | Centre and Littoral | Hospital | Non-probalistic | Women with pre/cancerous genital lesion | 29-87 | 52 IQR NR | Type specific PCR | Low | 175/181 = 96.69<br>NR |
| Sosso,2012 [19] | 2012 | Cross-sectional | Centre | Hospital | Non-probalistic | Sexually active non-pregnant women | 20-75 | 37.0 SD 3.0 | Abbott RealTime High-Risk HPV assay | Low | 107/278 = 38.49<br>80/107 = 74.77 |
| Teubeu,2014 [20] | 2012 | Cross-sectional | Centre and South-West | Community | Non-probalistic | Women | 30-65 | 41 IQR 36-50 | Abbott RealTime High-Risk HPV assay | Low | 146/529 = 27.60<br>NR |
| Bigoni,2013 [21] | 2013 | RCT | Centre | Community | Non-probalistic | Non-pregnant women | 25-65 | 41.4 SD 10.1 | Roche Cobas 4800 System | Low | 259/673 = 38.48<br>28/169 = 16.57 |
| Kabeyene,2014 [22] | 2014 | Cross-sectional | Centre | Hospital | Non-probalistic | Sexually active WLWH | 19-79 | 38.5 SD 9.9 | Abbott RealTime High-Risk HPV assay | Moderate | 10/17 = 58.82<br>NR |
| Akaaboune,2015 [23] | 2015 | Cross-sectional | West | Hospital | Non-probalistic | Non-pregnant women | 30-59 | 38.7 SD 5.6 | Cepheid GeneXpert | Low | 187/1012 = 18.48<br>10/187 = 5.35 |
| Domgue,2016 [24] | 2016 | Cross-sectional | North-West | Community | Non-probalistic | Non-pregnant women | 30-65 | 44.7 SD 9.8 | CareHPV | Low | 196/1270 = 15.43<br>16/136 = 11.76 |
| Fogue,2016 [25] | 2016 | Cross-sectional | West | Hospital | Non-probalistic | Sexually active women | 16-50 | 28.1 SD 8.0 | Type specific PCR | Low | 8/206 = 3.88<br>NR |
| Sosso,2016 [26] | 2016 | Cross-sectional | Centre | Hospital | Non-probalistic | Sexually active non-pregnant women | 18+ | 37.0 SD 6.5 | Abbott RealTime High-Risk HPV assay | Low | 73/257 = 28.40<br>55/73 = 75.34 |
| Cholli,2016 [27] | 2016 | Cross-sectional | Littoral and North-West | Hospital and community | Non-probalistic | Non-pregnant women | 30-80 | 41.8 SD 9.9 | CareHPV | Low | 223/913 = 24.42<br>157/223 = 70.40 |
| Essome,2017 [28] | 2017 | Cross-sectional | Littoral | Hospital | Non-probalistic | Women | 21-71 | 40.1 SD 10.0 | Indirect immunoperoxidase technique (P16INK4a) | Low | 44/108 = 40.74<br>NR |
| Rettig,2017 [29] | 2017 | Cross-sectional | North-West | Hospital | Non-probalistic | PLHIV | 24-69 | 42 IQR 37-48 | OraRisk (PCR-based) | Low | 4/101 = 3.96<br>NR |
| Castle,2018 [30] | 2018 | Cross-sectional | South-West | Hospital | Non-probalistic | WLWH | 25-59 | 42 IQR NR | Cepheid GeneXpert | Low | 257/486 = 52.88<br>NR |
| Doh,2018 [31] | 2018 | Cross-sectional | Centre | Hospital | Non-probalistic | Pregnant women | 25+ | NR | Roche Linear Array | Low | 25/464 = 5.39<br>33/62 = 53.23 |
| Levy,2018 [32] | 2018 | Cross-sectional | West | Community | Non-probalistic | Non-pregnant women | 30-49 | 39.4 SD 5.9 | Cepheid GeneXpert | Low | 166/840 = 19.76<br>10/166 = 6.02 |
| Wabo,2018 [33] | 2018 | Case-control | South-West | Hospital | Probabilistic | Women | 25-65 | 41.1 SD 10.5 | HPV High Risk Typing Real-TM | Low | 213/300 = 71.00<br>NR |
| Simo,2019 [34] | 2019 | Cross-sectional | Centre | Hospital | Non-probalistic | Women with pre/cancerous genital lesion | 20-84 | NR | Type specific PCR | Low | 370/410 = 90.24<br>102/370 = 27.57 |
| Vassilakos,2019 [11] | 2019 | Cross-sectional | West | Hospital | Probabilistic | Non-pregnant women | 30-50 | 40 IQR 35-45 | Abbott RealTime High-Risk HPV assay | Low | 66/1582 = 4.17<br>19/294 = 6.46 |
| Adedimeji,2020 [6] | 2020 | Cross-sectional | South-West | Hospital | Non-probalistic | Women | 19-59 | NR | Cepheid GeneXpert | Low | 324/855 = 37.89<br>260/324 = 80.25 |
| Broquet,2020 [35] | 2020 | Cross-sectional | West | Community | Non-probalistic | Non-pregnant women | 30-49 | 40 IQR 35-45 | Cepheid GeneXpert | Low | 382/2104 = 18.16<br>26/373 = 6.97 |
| Frund,2020 [36] | 2020 | Cross-sectional | West | Community | Non-probalistic | Women | NR | 38.3 IQR 27-46 | Cepheid GeneXpert | Low | 283/1606 = 17.62<br>NR |
| Manga,2020 [7] | 2020 | Cross-sectional | West, Centre and Littoral | Hospital | Non-probalistic | Female sex worker | 30+ | NR | AmpFire Assay | Low | 372/599 = 62.10<br>218/372 = 58.60 |
| Ngom,2020 [37] | 2020 | Cross-sectional | Centre and Littoral | Hospital | Non-probalistic | Patient with head and neck cancers | 20-78 | 49.0 SD NR | Abbott RealTime High-Risk HPV Assay | Low | 13/48 = 27.08<br>NR |
| Pham,2020 [38] | 2020 | Cross-sectional | West | Community | Non-probalistic | Non-pregnant women | 30-49 | 40.2 SD 5.9 | Cepheid GeneXpert | Low | 83/1940 = 4.28<br>NR |
| Simo,2020 [39] | 2020 | Cross-sectional | Centre | Hospital | Non-probalistic | Women | 20-65 | NR | Type specific PCR | Low | 36/135 = 26.67<br>NR |
| Mbimenyuy,2021 [40] | 2021 | Cross-sectional | Centre | Hospital | Non-probalistic | Sexually active non-pregnant women | 19+ | 44.0 SD 10.8 | Roche Linear Array | Low | 55/226 = 24.34<br>21/77 = 27.27 |
| Simo,2021 [41] | 2021 | Cross-sectional | Littoral | Hospital | Non-probalistic | Sexually active women | 20-70 | NR | Abbott RealTime High-Risk HPV assay | Low | 35/196 = 17.86<br>NR |
| Tchouaket,2021 [5] | 2021 | Cross-sectional | Centre and Littoral | Hospital | Non-probalistic | Sexually active non-pregnant women | 18+ | 41 IQR 34-50 | Abbott RealTime High-Risk HPV assay | Low | 78/364 = 21.43<br>2/364 = 0.55 |
| Enyegue,2023 [42] | 2023 | Cross-sectional | Centre | Hospital | Non-probalistic | Sexually active women | 15-65 | NR | QIAGEN Multiplex PCR Kit | Low | 27/343 = 7.87<br>65/105 = 61.90 |
| Manga,2023 [43] | 2023 | Cross-sectional | Littoral | Community | Non-probalistic | WSW | 19-40 | 29.2 SD 5.3 | AmpFire Assay | Low | 11/19 = 57.89<br>0/11 = 0.00 |

1: Year of study completion; 2: Prevalence of high-risk human papillomavirus (HPV) infections among screened participants; 3: Prevalence of human immunodeficiency virus (HIV) infections among screened HPV positive participants; RCT: Randomized control trial; WLWH: Women living with HIV; PLVIH; People living with HIV; NR: Not reported; IQR: Interquartile range; SD: Standard deviation; WSW: Women having sex with women; PCR: Polymerase chain reaction;

### 3.3. High-risk human papillomavirus

The pooled prevalence of high-risk HPV was 28.95 (95%CI: 19.74-40.30; 33 studies; n = 18,798 participants) with high heterogeneity observed across studies (*I*^2^ = 98.7%; p D 0.001) (**Fig. 2**).

**Fig. 2.**
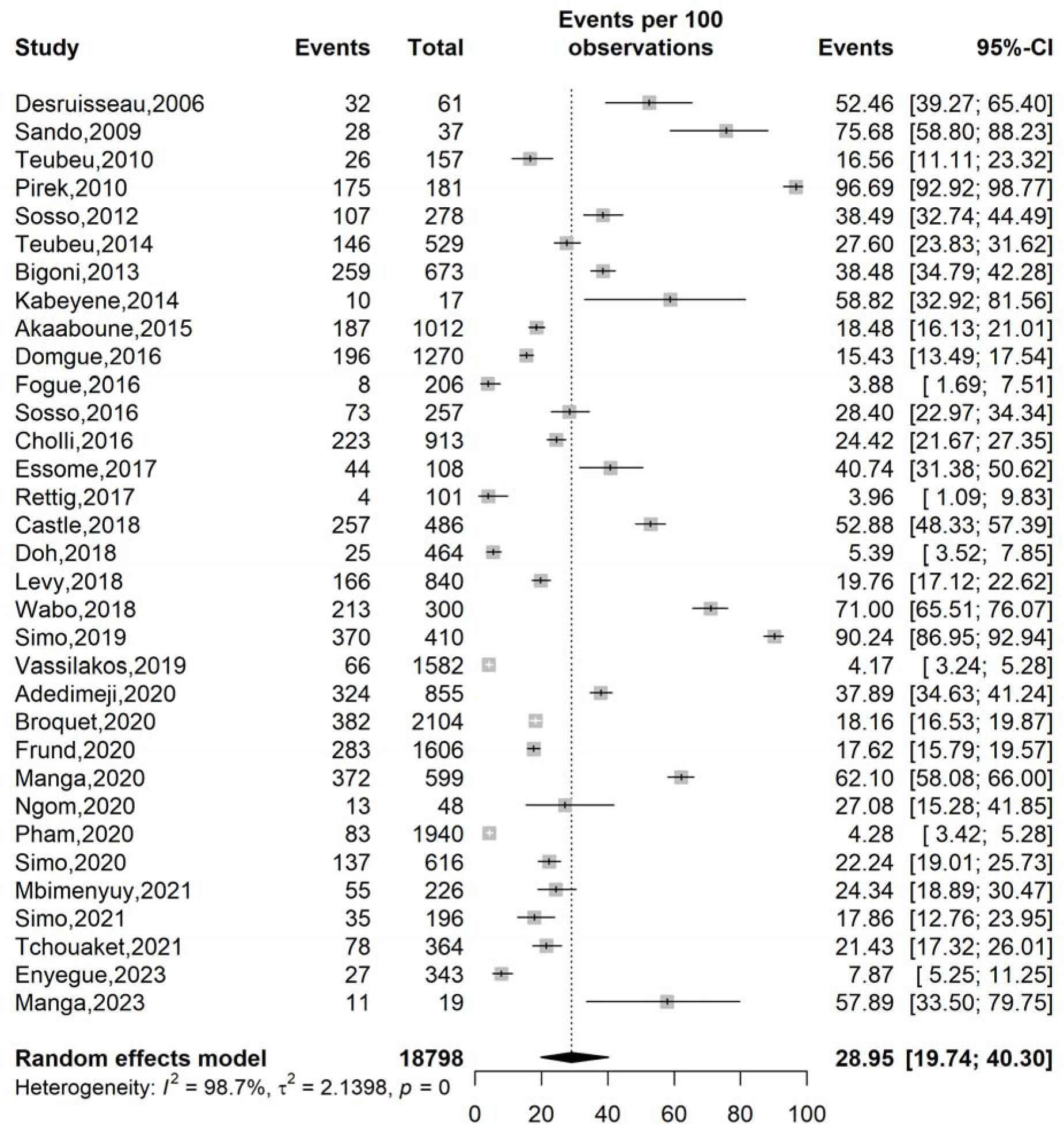
Prevalence of high-risk human papillomavirus infections in Cameroon

There was a decline in high-risk HPV prevalence over time, from 53.34% before 2014 (26.98-77.96; 7 studies; 1,916 participants) to 21.43% (95%CI: 11.81-35.82; 5 studies; n = 1,148 participants) in 2021–2023.

The study revealed that pooled high-risk HPV prevalence was high in multi-region studies conducted in Centre and Littoral regions (58.71; 95%CI: 10.99-94.24; 3 studies; n = 593 participants), and in single region studies conducted in the South-West (54.12%; 95%CI: 3 studies; n = 1,641 participants), although observed in a single studies, one study conducted in the Centre, Littoral and West regions presented a high prevalence of 62.10% (95%CI : 58.08-66.00; n = 599 participants). Conversely, the lowest prevalences were observed in the North-West (8.89%; 95%CI: 3.26-22.03; 2 studies; n = 1,371 participants) and West regions (10.79%; 95%CI:6.53-17.31; 8 studies; n = 9,447 participants).

Although the subgroup difference was not statistically significant (p = 0.092). A significantly higher prevalence (p = 0.033) of HR-HPV was observed in other studies design including cohort and randomized control trial studies (55.6%; 95%CI: 32.28-76.05; 2 studies; n = 973 participants as compared to cross-sectional studies (27.47%; 95CI: 18.32-39.02; 31 studies; 17,825 participants). The highest prevalences were significantly (*p* D 0.001) observed among women with pre/cancerous lesions (90.69%; 95%CI:78.48-96.30; 3 studies; n = 628 participants) and female sex workers (62.10%; 95%CI: 58.08-66.00; 1 study; 599 participants) as compared to women from general population (21.09%; 95%CI: 15.16-28.55; 25 studies; n = 16,919 participants) (**Table 2 and Additional file 1, Supplementary Fig. 1-7**)

**Table 2.** Subgroup meta-analysis of the pooled prevalence of high-risk human papillomavirus infection in Cameroon.

| Subgroup | Sample size | Prevalence (%) | 95% CI limits<sup>1</sup> |  | Number of studies | Heterogeneity statistic<sup>1</sup> |  | Subgroup difference p-value |
| --- | --- | --- | --- | --- | --- | --- | --- | --- |
|  |  |  | Lower | Upper |  | I<sup>2</sup>(%) | p-value |  |
| **Study period (year)<sup>2</sup>** |  |  |  |  |  |  |  | 0.092 |
| □ 2014 | 1,916 | 53.34 | 26.98 | 77.96 | 7 | 96.2 | □ 0.001 |  |
| 2014-2020 | 15,734 | 23.96 | 14.57 | 36.78 | 21 | 99.1 | □ 0.001 |  |
| 2021-2023 | 1,148 | 21.43 | 11.81 | 35.82 | 5 | 91.2 | □ 0.001 |  |
| **Study design** |  |  |  |  |  |  |  | 0.033 |
| Cross-sectional | 17,825 | 27.47 | 18.32 | 39.02 | 31 | 98.6 | □ 0.001 |  |
| Other | 973 | 55.16 | 32.28 | 76.05 | 2 | 98.8 | □ 0.001 |  |
| **Study setting** |  |  |  |  |  |  |  | 0.414 |
| Community | 8,981 | 20.72 | 12.40 | 32.55 | 8 | 98.4 | □ 0.001 |  |
| Hospital | 8,904 | 32.15 | 19.76 | 47.69 | 24 | 98.7 | □ 0.001 |  |
| Hospital & community | 913 | 24.42 | 21.67 | 27.35 | 1 | - | - |  |
| **Region** |  |  |  |  |  |  |  | □ 0.001 |
| Centre | 3,321 | 35.41 | 18.04 | 57.73 | 10 | 98.4 | □ 0.001 |  |
| Littoral | 323 | 35.14 | 18.73 | 73.89 | 3 | 92.1 | □ 0.001 |  |
| North-West | 1,371 | 8.89 | 3.26 | 22.03 | 2 | 88.0 | □ 0.001 |  |
| South-West | 1,641 | 54.12 | 38.28 | 69.17 | 3 | 98.0 | □ 0.001 |  |
| West | 9,447 | 10.79 | 6.53 | 17.31 | 8 | 97.9 | □ 0.001 |  |
| Centre & Littoral | 593 | 58.71 | 10.99 | 94.24 | 3 | 98.3 | □ 0.001 |  |
| Centre & South-West | 590 | 38.05 | 22.68 | 56.25 | 2 | 93.3 | □ 0.001 |  |
| Centre, Littoral, & West | 599 | 62.10 | 58.08 | 66.00 | 1 | - | - |  |
| Littoral & North-West | 323 | 35.14 | 18.73 | 56.00 | 1 | - | - |  |
| Multi-region<sup>3</sup> | 2,695 | 47.99 | 23.13 | 73.89 | 7 | 98.3 | □ 0.001 |  |
| **Sampling method** |  |  |  |  |  |  |  | 0.870 |
| Non-probabilistic | 16,916 | 29.24 | 19.97 | 40.63 | 31 | 98.5 | □ 0.001 |  |
| Probabilistic | 1,882 | 24.58 | 1.94 | 84.27 | 2 | 99.8 | □ 0.001 |  |
| **Participant** |  |  |  |  |  |  |  | □ 0.001 |
| Women from general population | 16,919 | 21.09 | 15.16 | 28.55 | 25 | 98.2 | □ 0.001 |  |
| PLHIV | 604 | 28.88 | 5.98 | 72.14 | 3 | 95.1 | □ 0.001 |  |
| Women with pre/cancerous genital lesions | 628 | 90.69 | 78.48 | 96.30 | 3 | 87.3 | □ 0.001 |  |
| Patient with head & neck cancers | 48 | 27.08 | 15.28 | 41.85 | 1 | - | - |  |
| Female sex worker | 599 | 62.10 | 58.08 | 66.00 | 1 | - | - |  |

<sup>1</sup> Random effects model; <sup>2</sup> □ 2014 = Pre-HPV vaccination in Cameroon, 2014-2020= HPV vaccination demonstration in Cameroon, 2021-2023= post-HPV vaccine introduction in Cameroon; <sup>3</sup> Multi-region included studies conducted in more than one of following regions (Centre, Littoral, South-West, North-West and West); CI: Confidence interval; PLHIV: People living with human immunodeficiency virus (HIV)

### 3.4. HIV and human papillomavirus

The pooled prevalence of HPV among HPV-positive individuals was 26.17 (95%CI: 13.48-44.65; 19 studies; n = 3,589 participants) with high heterogeneity observed between studies (*I*^2^ = 97.9%; *p* D 0.001) (**Fig. 3**).

**Fig. 3.**
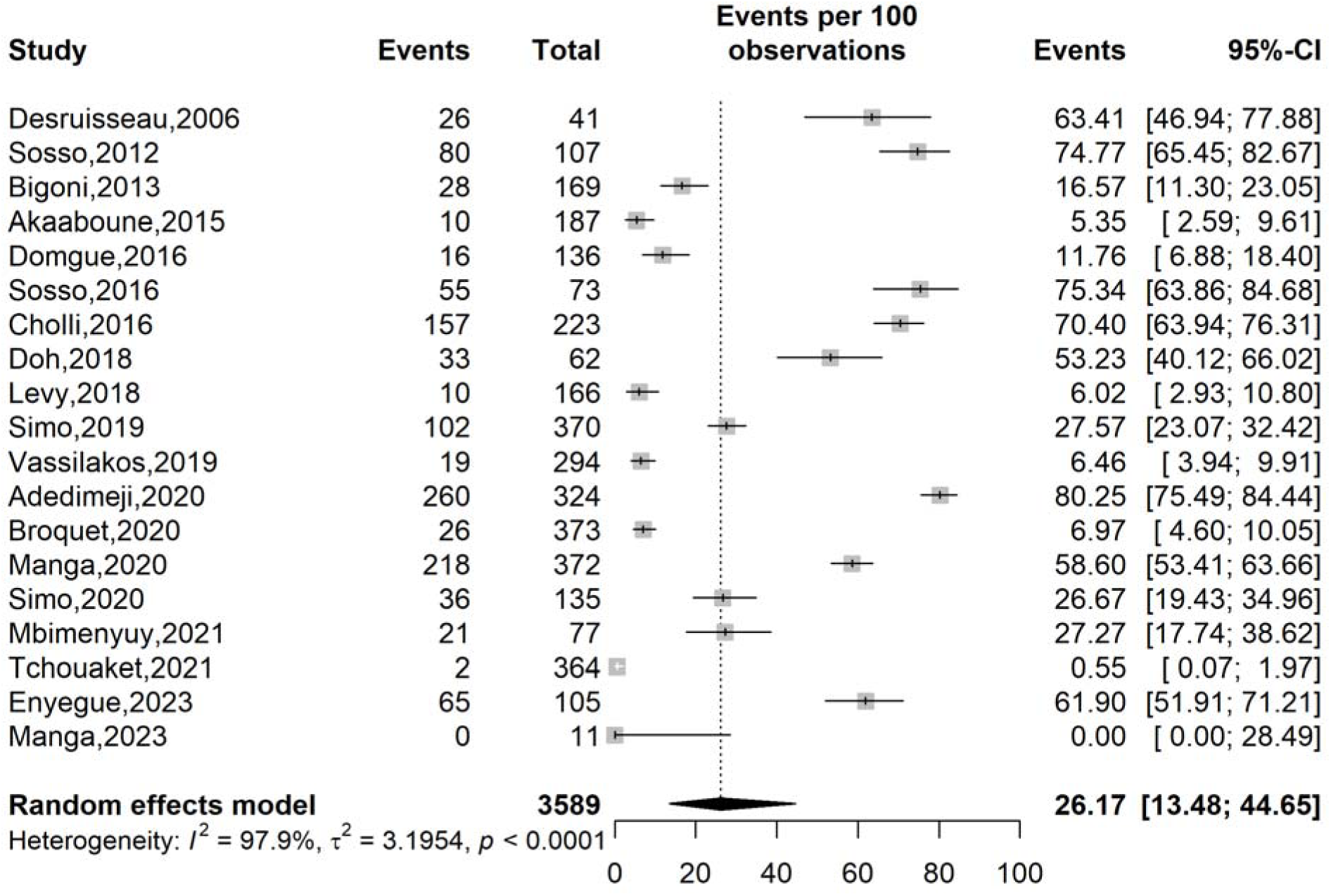
Prevalence of HIV infections among human papillomavirus-positive cases in Cameroon

Although non-statistically significant (*p* = 0.223), we observed a progressive decline of HIV prevalence among HPV-positive cases in Cameroon from 50.01% (95%CI: 20.57-79.44; 3 studies; n = 317 participants) before 2014 to 7.02% (95%CI: 0.47-54.56; 4 studies; n = 557 participants).

Studies exclusively conducted in hospitals revealed significantly (*p* D 0.001) higher prevalence (34.57%; 95%CI: 16.42-58.71; 13 studies; n = 2,511 participants) than community-based studies (9.18%; 95%CI: 6.02-13.76; 5 studies; n = 855 participants).

Among single region-based studies, the pooled prevalence was significantly (*p* D 0.001) higher in the Centre region (44.32%; 95%CI: 28.52-61-36; 8 studies; n = 1,098 participants) compared to the West region (6.37%; 95%CI: 5.03-8.05; 4 studies; n = 1,020 participants).

Although evidenced in a single study, the prevalence of HIV among HPV-positive individuals were significantly (*p* D 0.001) higher among female sex workers (58.60%; 95%CI: 23.07-32.42; 370 participants) than those from general population (24.34%; 95%CI: 11.43-44.51; 17 studies; n = 2,847 participants) (**Table 3 and Additional file 1, Supplementary Fig.10-16)**

**Table 3.** Subgroup meta-analysis of the pooled prevalence of HIV infections among human papillomavirus-positive women in Cameroon.

| Subgroup | Sample size | Prevalence (%) | 95% CI limits<sup>1</sup> |  | Number of studies | Heterogeneity statistic<sup>1</sup> |  | Subgroup difference |
| --- | --- | --- | --- | --- | --- | --- | --- | --- |
|  |  |  | Lower | Upper |  | I<sup>2</sup>(%) | p-value |  |
| **Study period (year)<sup>2</sup>** |  |  |  |  |  |  |  | 0.223 |
| □ 2014 | 317 | 50.01 | 20.57 | 79.44 | 3 | 97.7 | □ 0.001 |  |
| 2014-2020 | 2,175 | 28.53 | 14.00 | 49.48 | 12 | 98.5 | □ 0.001 |  |
| 2021-2023 | 557 | 7.02 | 0.47 | 54.55 | 4 | 95.7 | □ 0.001 |  |
| **Study design** |  |  |  |  |  |  |  | 0.213 |
| Cross-sectional | 3,420 | 26.75 | 13.28 | 46.54 | 18 | 97.9 | □ 0.001 |  |
| Randomized control trial | 169 | 16.57 | 11.30 | 23.05 | 1 | - | - |  |
| **Study setting** |  |  |  |  |  |  |  | □ 0.001 |
| Community | 855 | 9.18 | 6.02 | 13.76 | 5 | 97.7 | 0.005 |  |
| Hospital | 2,511 | 34.57 | 16.42 | 58.71 | 13 | 97.7 | □ 0.001 |  |
| Hospital & community | 223 | 70.40 | 63.94 | 76.31 | 1 | - | - |  |
| **Region** |  |  |  |  |  |  |  | □ 0.001 |
| Centre | 1,098 | 44.32 | 28.52 | 61.36 | 8 | 95.9 | □ 0.001 |  |
| Littoral | 11 | 0.00 | 0.00 | 28.49 | 1 | - | - |  |
| North-West | 136 | 11.76 | 6.88 | 18.40 | 1 | - | - |  |
| South-West | 324 | 80.25 | 75.49 | 84.44 | 1 | - | - |  |
| West | 1,020 | 6.37 | 5.03 | 8.05 | 4 | 0.0 | 0.899 |  |
| Centre & Littoral | 364 | 0.55 | 0.07 | 1.97 | 1 | - | - |  |
| Centre & South-West | 41 | 63.41 | 46.94 | 77.88 | 1 | - | - |  |
| Centre, Littoral, & West | 372 | 58.60 | 53.41 | 63.66 | 1 | - | - |  |
| Littoral & North-West | 223 | 70.40 | 63.94 | 76.31 | 1 | - | - |  |
| Multi-region<sup>3</sup> | 1,000 | 30.30 | 3.55 | 83.69 | 4 | 95.9 | □ 0.001 |  |
| **Sampling method** |  |  |  |  |  |  |  | 0.870 |
| Non-probabilistic | 3,295 | 28.04 | 14.32 | 47.59 | 18 | 97.8 | □ 0.001 |  |
| Probabilistic | 294 | 6.46 | 3.94 | 9.91 | 1 | - | - |  |
| **Participant** |  |  |  |  |  |  |  | □ 0.001 |
| Women from general population | 2,847 | 24.34 | 11.43 | 44.51 | 17 | 98.0 | □ 0.001 |  |
| Women with pre/cancerous genital lesions | 370 | 27.57 | 23.07 | 32.42 | 1 | - | - |  |
| Female sex worker | 372 | 58.60 | 53.41 | 63.66 | 1 | - | - |  |

<sup>1</sup> Random effects model; <sup>2</sup> □ 2014 = Pre-HPV vaccination in Cameroon, 2014-2020= HPV vaccination demonstration in Cameroon, 2021-2023= post-HPV vaccine introduction in Cameroon; <sup>3</sup> Multi-region included studies conducted in more than one of following regions (Centre, Littoral, South-West, North-West, and West); CI: Confidence interval; PLHIV: People living with human immunodeficiency virus (HIV)

### 3.5. Genotype profile of human papillomavirus among affected women

Most high-risk HPV strains identified among HPV-positive women belonged to the serotype 16 (28.7%), 52(23.6%), 66(22.9%) and 33(22.8%), the HPV 18 mostly responsible for uterine cervical cancer along with HPV 16 presented a slightly lower prevalence around 20.2% (**Fig. 4**). Most HPV coinfection cases included HPV 39/56/66/68 (31.2%), HPV 51/59 (18.5%), 16/18/45 (11.5%), and 18/45 (11.2%) (**Additional file 2; Supplementary Fig. 1**).

**Fig. 4.**
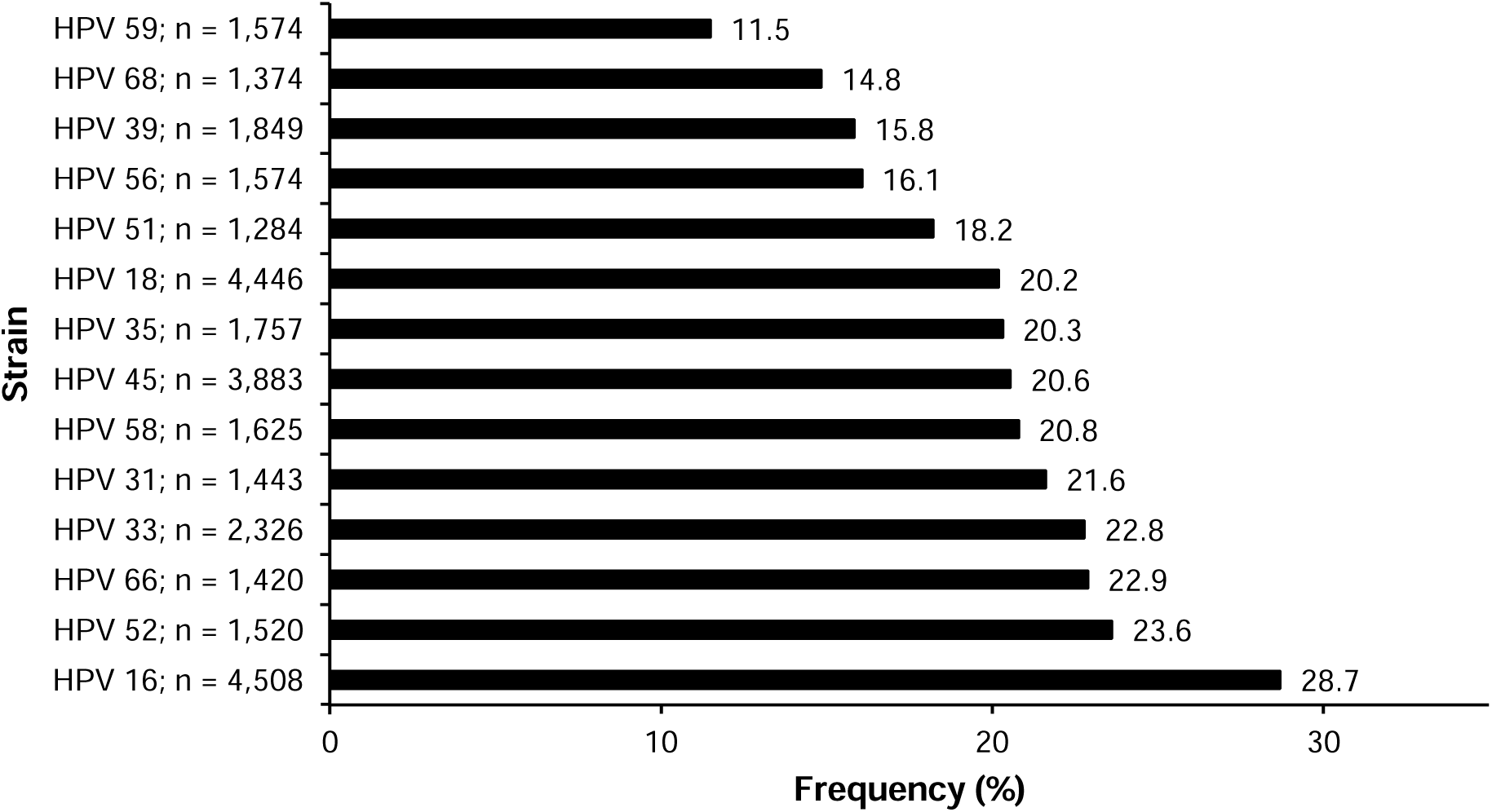
Trend of human papillomavirus genotypes among human papillomavirus-positive women in Cameroon

### 3.6. Abnormal cervical lesions among human papillomavirus-positive women

A total of 12 studies identified 637 abnormal cervical lesions among 2,186 HPV-positive women resulting in a pooled prevalence of 35.15% (95%CI: 20.21-53.70; I2 = 95.2%; *p* 0.001) (**Fig. 5**).

**Fig. 5.**
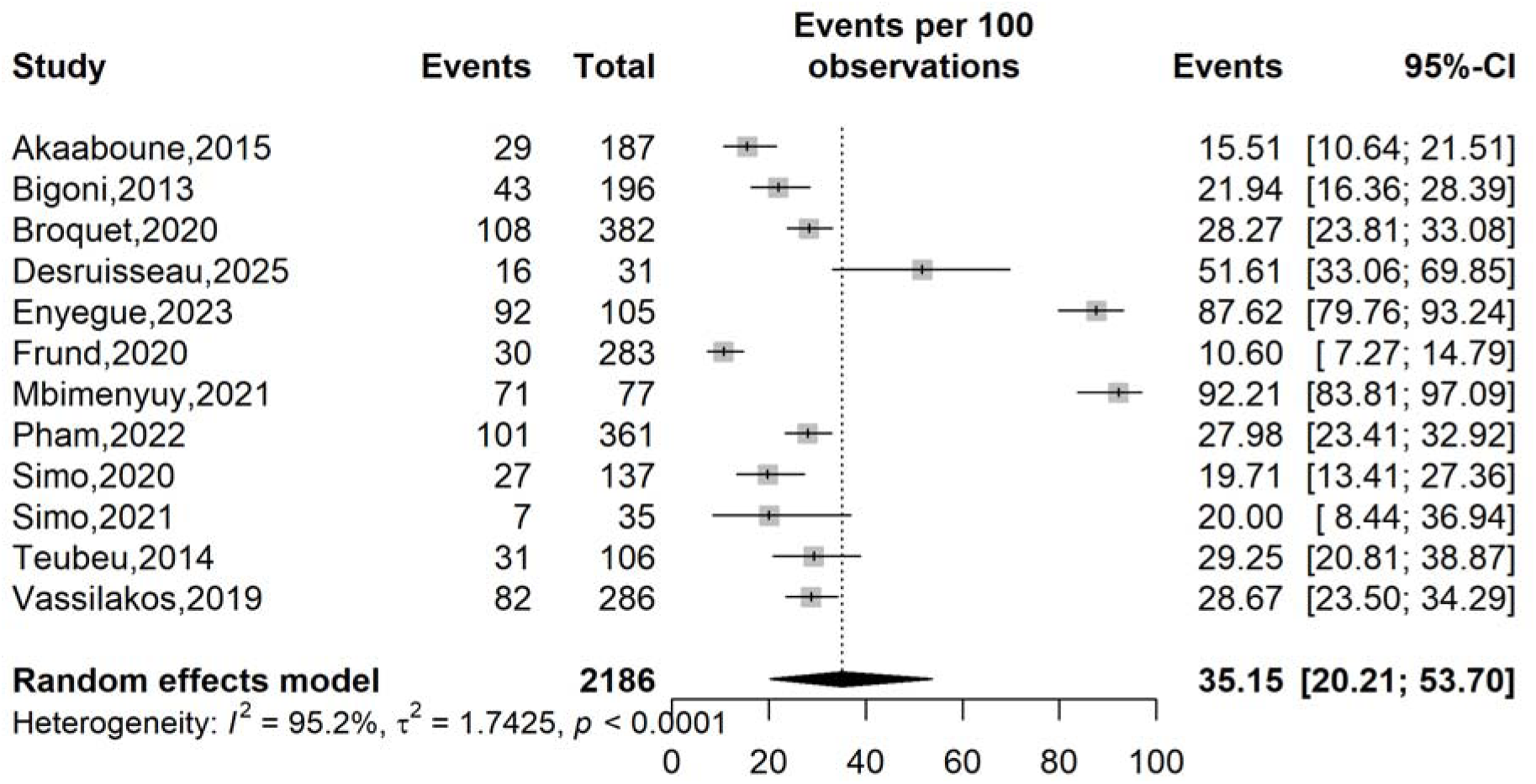
Pooled prevalence of abnormal uterine cervical lesions among human papillomavirus-positive women in Cameroon

The most commonly observed uterine cervical lesions among HPV-positive women included any cervical lesions (38.4%; 95%CI: 28.0-49.9; 1 study; n = 73 participants), followed by low-grade squamous intra-epithelial lesions (LSIL = 34.4%; 95%CI: 51.1-58.1; 7 studies; n = 765 participants), high-grade squamous intra-epithelial lesions (HSIL = 19.1%; 95%CI: 21.3-27.3; 8 studies; n = 785 participants); and Intraepithelial cervical carcinomas (ICC = 18.2%; 95%CI: 11.1-28.4; 1 study; 77 participants) (**Fig. 6 and Additional file 2, Supplementary Fig. 2**).

**Fig. 6.**
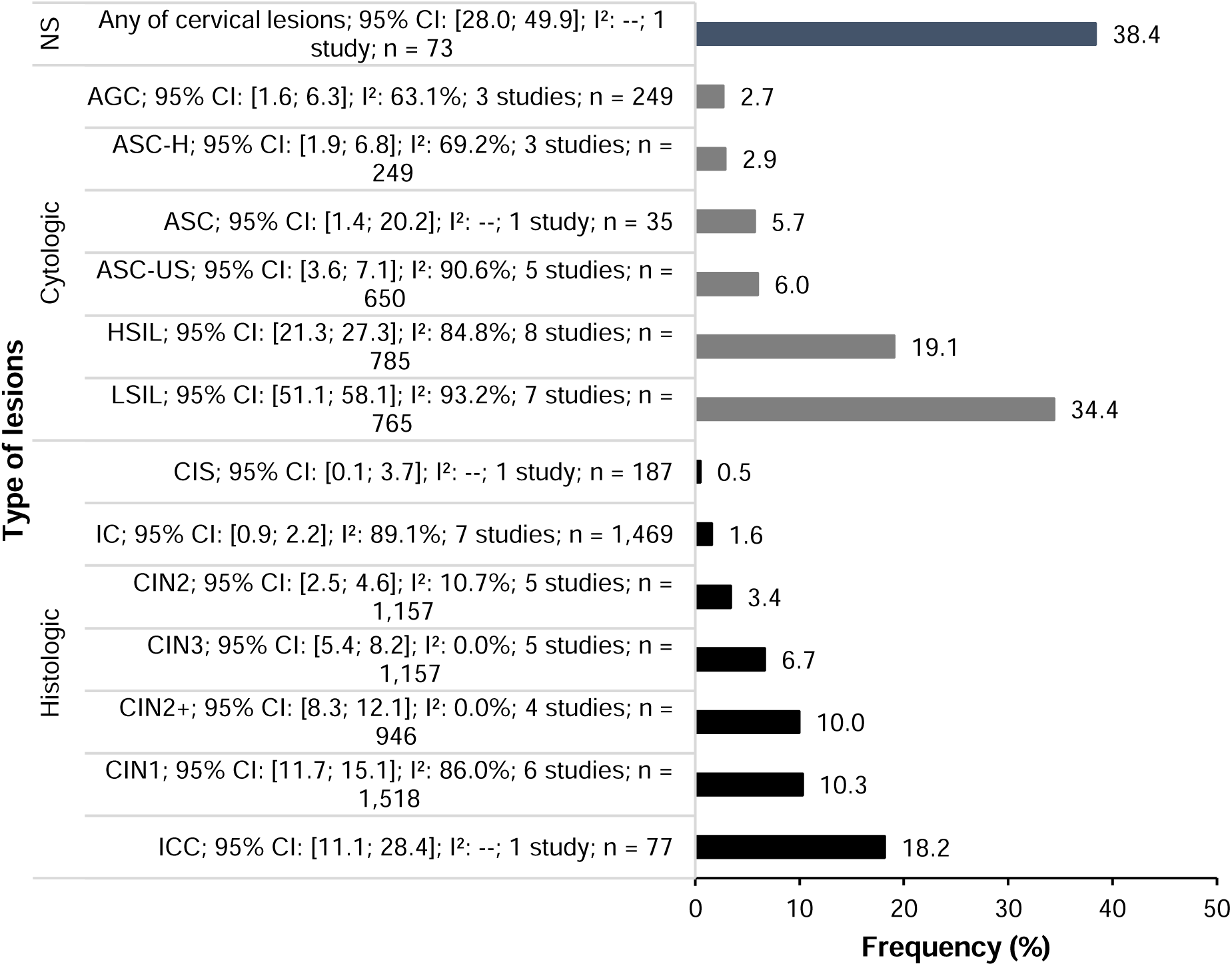
Uterine cervical lesions observed among human papillomavirus-positive women in Cameroon. (**CIS** = Carcinoma in situ, **IC**= Invasive carcinoma; **AGC**=Atypical glandular cells; **ASC-H**=Atypical squamous cells, cannot exclude high-grade squamous intraepithelial lesion; **CIN2**=Cervical intraepithelial neoplasia grade 2; **ASC**=Atypical squamous cells; **ASC-US**=Atypical squamous cells of undetermined significance; **CIN3**=Cervical intraepithelial neoplasia grade 3; **CIN2+**= Cervical intraepithelial neoplasia grade 2 or more; **CIN1**=Cervical intraepithelial neoplasia grade 1; **ICC**=Intraepithelial cervical carcinoma; **HSIL**=High-grade squamous intraepithelial lesion, includes CIN2 and CIN3; **LSIL**=Low-grade squamous intraepithelial lesion, includes CIN1 and mild HPV changes; **Any of cervical lesions**=A combined category representing any abnormal cervical findings; **NS** = Not specified.)

Following the group of lesions categorized as any cervical lesions, the most commonly identified category included low-grade lesions (19.8%; 95%CI: 10.59-33.95; 13 studies; n = 2,283 participants), high-grade lesions (15.6%; 95%CI: 9.98-23.69; 14 studies; n = 2,586 participants); and Atypical Squamous lesions (8.8%; 95%CI: 3.06-22.81; n = 650 participants) (**Fig. 7 and Additional file 2, Supplementary Fig. 3**).

**Fig. 7.**
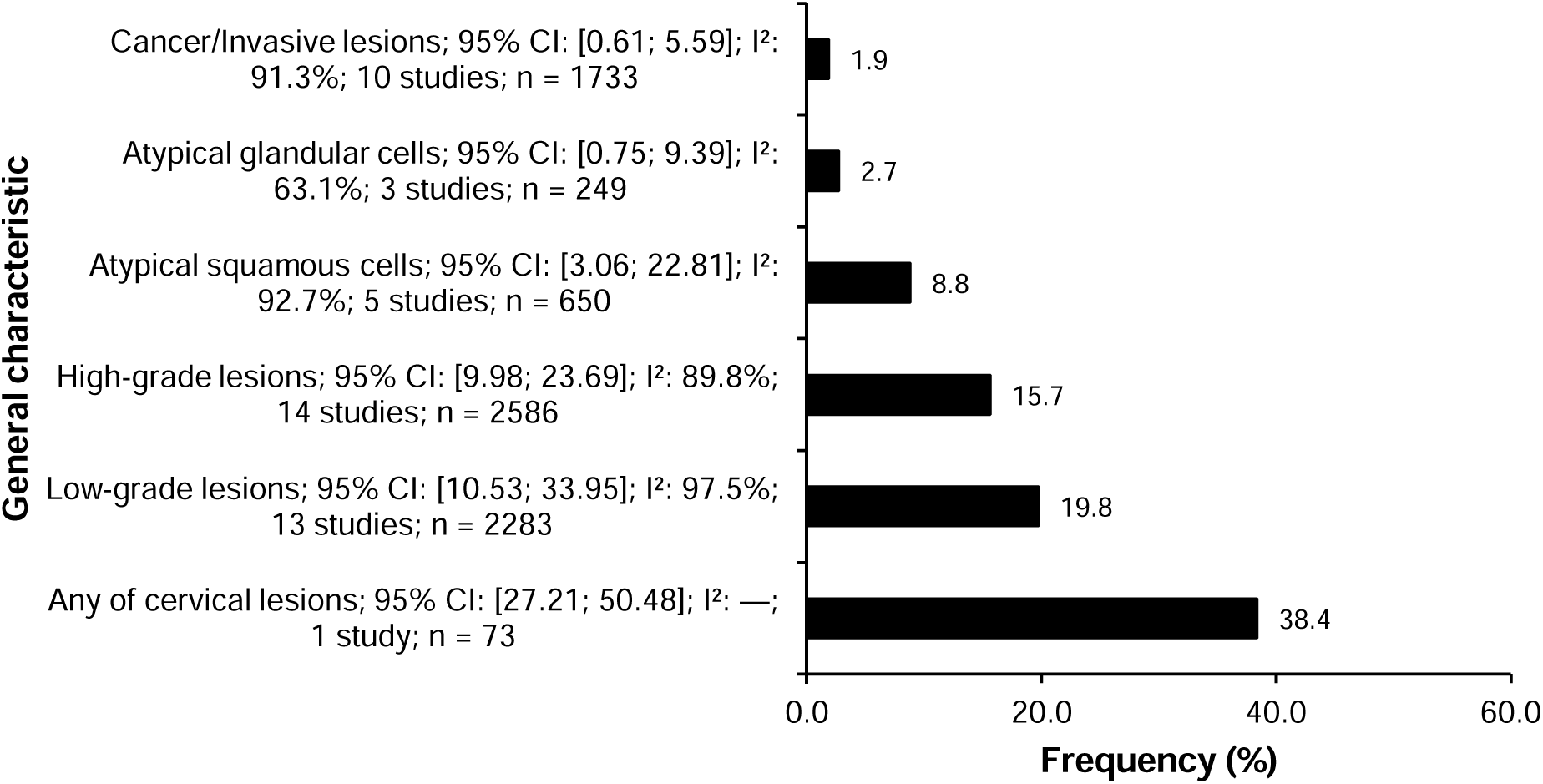
Uterine cervical lesion categories observed among human papillomavirus-positive women in Cameroon. (**ASC** = Atypical squamous cells + undetermined significance + cannot exclude high-grade; **Low-grade** = Low-grade squamous intraepithelial lesion + cervical intraepithelial neoplasia grade 1; **High-grade** = Low-grade squamous intraepithelial lesion + cervical intraepithelial neoplasia grade 2 and more; **Cancer/Invasive lesions** = Carcinoma in situ, Intraepithelial cancer, and Invasive cancer)

### 3.7. Publication bias and sensitivity analysis

Despite the asymmetric funnel plot observed, the Egger’s (*p* = 0.570) and Begg’s (*p* = 0.495) tests revealed non-significant risk of publication bias for the HR HPV pooled prevalence (**Additional file 1, Supplementary Fig. 9**). The sensitivity analysis showed no study significantly influenced the overall pooled HR HPV infection prevalence estimate (**Additional file 1, Supplementary Fig. 8**). Similar observations were made for the HIV prevalence among HPV-positive individuals where non-significant risk of publication bias was identified (Egger’s (*p* = 0.070) and Begg’s (*p* = 0.100) tests) and no single study significantly influenced the resulting pooled estimate (**Additional file 1, Supplementary Fig. 17 and 18**).

## 4. Discussion

This systematic review and meta-analysis present the first comprehensive national overview of HR-HPV prevalence in Cameroon, including the burden of HIV co-infection and abnormal cervical lesions among HPV-positive women. By combining data from 33 studies involving 18,798 participants, our findings offer critical baseline evidence in the context of ongoing HPV vaccination and cervical cancer prevention efforts in the country.

Our review found that the pooled prevalence of HR-HPV in Cameroon was 28.95% (95% CI: 19.74–40.30; 33 studies; n=18,798), with very high heterogeneity. This prevalence is considerably higher than global averages reported in many high-income settings [44]. However, it aligns with the pooled HR-HPV prevalence reported for sub-Saharan Africa, where a recent systematic review and meta-analysis identified an overall pooled prevalence of about 34% and similarly high heterogeneity across countries and study designs [45]. This similarity stems from shared epidemiologic factors in sub-Saharan Africa, such as high background HPV exposure, limited screening, significant burden of HIV, poor HPV vaccine coverage. Similarly, we observed a higher prevalence of HR-HPV in facility-based studies (34.57%) than in community-based studies (9.18%). This could be due to selection bias. Women who go to health facilities are more likely to have symptoms, be referred for abnormal results, or have HIV. These factors are associated with higher rates and greater persistence of HR-HPV. This pattern has been observed elsewhere in sub-Saharan Africa [46].

Also, we observed a more than 50% decline in HR-HPV prevalence, from 53.34% before 2014 to 21.43% between 2021 and 2023. While this trend is encouraging, it should be viewed with caution, as these numbers come from studies with different populations and methods, so the decrease may be due to changes in testing, sampling, or the risk profiles of the groups studied, rather than a true cause-and-effect relationship. Since national HPV vaccination was effective only from 2020, most adult women in these studies would not yet have shown a clear reduction in HR-HPV rates due to the vaccine [47]. To properly measure the vaccine’s impact, age-specific monitoring and repeated surveys in groups eligible for vaccination will be needed. Furthermore, this could also be explained by increased health promotion activities on HPV rather than by the vaccines.

There was also a significant regional variation in pooled HR-HPV prevalence across Cameroon in our analysis, with higher rates recorded in the South-West and some multi-region studies, and lower rates in the West and North-West. Similar differences within countries have been widely reported across sub-Saharan Africa. These are often linked to factors such as differences in HIV rates, the structure of urban and rural sexual networks, access to screening, care-seeking behaviors, and disruptions to health systems caused by conflict [45, 46]. In Cameroon, most studies were conducted in the Centre and Littoral regions, which are mainly urban referral settings. This focus may lead to higher pooled ‘national’ estimates if higher-risk groups are overrepresented.

Further in our analysis, we observed a high rate of HR-HPV among women with pre-cancerous or cancerous lesions (90.69%), and the elevated rate among female sex workers (62.10%) in Cameroon. Our results are consistent with previous studies across sub-Saharan Africa, which showed that HR-HPV infection is almost universal in women on the cervical neoplasia pathway and is especially common in high-risk sexual networks. Regional meta-analyses also report very high HPV detection in women with CIN2+ and invasive cervical cancer, as well as higher rates of HPV and multiple infections among female sex workers, likely due to greater exposure and reinfection [48, 49].

In addition, our meta-analysis found that 26.17% of HPV-positive women also had HIV, with higher rates in facility-based studies than in community-based ones. This likely reflects both the known link between HIV and HPV in sub-Saharan Africa and the fact that more women living with HIV are seen in hospitals and referral centers. Other regional studies show that women living with HIV have higher rates of HPV infection, persistence, and multiple HPV types than HIV-negative women [50]. Based on this, the World Health Organization recommends that women living with HIV start cervical cancer screening at age 25 and get screened more often using high-performance HPV DNA tests [51]. Our results support integrating HPV-based screening into HIV care in Cameroon as an effective prevention strategy. The most common HR-HPV genotypes among HPV-positive women in our synthesis were HPV 16 (28.7%), followed by HPV 52 (23.6%), HPV 66 (22.9%), HPV 33 (22.8%), and HPV 18 (20.2%). This pattern is similar to what has been reported in sub-Saharan Africa, where HPV16 is most common and types like HPV52, HPV33, and HPV35 also play a major role in HR-HPV circulation and cervical cancer cases [52].

These results are important for vaccine planning. The nonavalent vaccine, which covers HPV 16, 18, 31, 33, 45, 52, and 58, offers broader protection against genotypes commonly found in Cameroon, especially HPV 33 and HPV 52, compared to the bivalent and quadrivalent vaccines which are currently in use in Cameroon [52]. Further feasibility studies should be conducted to introduce the nonvalent vaccine in Cameroon. Also, since HPV 66 was common in our study and is not included in current vaccines, it is important to keep monitoring which genotypes are present locally. This will help ensure that vaccination and screening programs remain effective.

We also observed in our study that in HPV-positive women, 38.4% had some form of cervical lesion. LSIL was found in 34.4%, HSIL in 19.1%, and intraepithelial cervical carcinoma in 18.2%. Grouped together, low-grade lesions made up 19.8%, high-grade lesions 15.6%, and atypical squamous lesions 8.8%. A higher number of low-grade lesions shows the early stage of HPV infection. However, the notable rates of high-grade lesions and carcinoma point to a significant burden of advanced precancerous cervical disease. Studies in sub-Saharan Africa have found similar trends, where ongoing HR-HPV infection is the main cause of cervical cancer [48].

## 5. Limitations

This review has some important limitations. The pooled analyses showed substantial heterogeneity due to differences in methods and populations. This could affect how accurate the summary estimates are. Most of the studies included were cross-sectional and hospital-based, which could introduce referral bias and overestimate HR-HPV prevalence compared with the general population. Although studies from different regions were included, the coverage was uneven, so the pooled estimates may mostly reflect urban and higher-risk areas. Many studies used non-probabilistic sampling, which raises the risk of selection bias. Limited age-specific data made it difficult to assess vaccine-eligible groups, and the lack of male data limits our understanding of transmission.

## 6. Conclusion

The overall high rate of HR-HPV, the significant number of precancerous and intraepithelial lesions in HPV-positive women, and the genotype patterns with clear vaccine implications all point to an ongoing risk of cervical cancer. While some regions remain underrepresented, these results show that it is urgent to expand HPV vaccination, assess the feasibility of introducing nonavalent vaccines in Cameroon to cover most HPV subtypes, make high-quality HPV screening more accessible to everyone, and improve early detection and treatment. Accelerating integrated prevention strategies will be essential for reducing cervical cancer burden in Cameroon and advancing progress toward global elimination targets.

## Supporting information

Addition file 1

## 7. Data Availability

All data generated or analyzed during this study are included in this published article and its supplementary information files

## 8. Abbreviations

*AGC*: Atypical glandular cells
*ASC*: Atypical squamous cells
*ASC-H*: Atypical squamous cells, cannot exclude high-grade squamous intraepithelial lesion
*ASC-US*: Atypical squamous cells of undetermined significance
*CI*: Confidence interval
*CIN1*: Cervical intraepithelial neoplasia grade 1
*CIN2*: Cervical intraepithelial neoplasia grade 2
*CIN2+*: Cervical intraepithelial neoplasia grade 2 or more
*CIN3*: Cervical intraepithelial neoplasia grade 3
*CIS*: Carcinoma in situ
*HIV*: Human immunodeficiency virus
*HPV*: Human papillomavirus
*HR-HPV*: High-risk human papillomavirus
*HSIL*: High-grade squamous intraepithelial lesion, includes CIN2 and CIN3
*IC*: Invasive carcinoma
*ICC*: Intraepithelial cervical carcinoma
*IQR*: Interquartile range
*LSIL*: Low-grade squamous intraepithelial lesion, includes CIN1 and mild HPV changes;
*MeSH*: Medical subject headings
*NR*: Not reported
*NS*: Not specified
*PCR*: Polymerase chain reaction
*PLHIV*: people living with HIV
*PRISMA*: Preferred reporting items for systematic reviews and meta-analysis
*RCT*: Randomized control trial *SD*: Standard deviation
*WLWH*: Women living with HIV
*WSW*: Women having sex with women

## 8. Declarations

### Ethical approval and consent to participate

Not applicable.

### Consent for publication

Not applicable.

### Competing interests

All authors declare no conflicts of interest and have approved the final version of the article.

### Funding source

This research did not receive any specific grant from funding agencies in the public, commercial or not-for-profit sectors.

### Author contributions

FZLC conceived the original idea of the study; FZLC and RT conducted the literature search; FZLC, RT, CA, LBKB, ELMB selected the studies, extracted the relevant information, and synthesized the data; FZLC performed the analyses; FZLC, LBKB, RT, and CA wrote the first draft of the manuscript. All authors critically reviewed and revised successive drafts of the manuscript. All authors read and approved the final manuscript.

## Acknowledgements

None.

## 9. Additional files

### Additional file 1

- Searching strategy by databases
- Subgroup analysis of HR-HPV prevalence in Cameroon
- Sensitivity analysis of HR-HPV prevalence in Cameroon
- Publication bias of HR-HPV prevalence in Cameroon
- Subgroup analysis of HIV co-infection prevalence in Cameroon
- Sensitivity analysis of HIV co-infection prevalence in Cameroon
- Publication bias of HIV co-infection prevalence in Cameroon
- Publication bias of abnormal cervical lesions prevalence in Camerron

### Additional file 2

- Trend of HPV strains identified in Cameroon
- Cervical lesions observed among HPV-positive women in Cameroon

## Notes

### Competing Interest Statement

The authors have declared no competing interest.

### Author Declarations

All data source are available in the reference list of the manuscript.

