## Additional file 2 for "Prevalence of high-risk human papillomavirus genotypes and associated HIV co-infection in Cameroon: a systematic review and meta-analysis"

**Genotypic profile**

| HPV strain | Frequency (%) |
| --- | --- |
| 39/56/66/68; 95% CI: [26.20; 36.55]; I²: —; 1 study; n = 324 | 31.17 |
| 51/59; 95% CI: [14.53; 23.29]; I²: —; 1 study; n = 324 | 18.52 |
| 16; 95% CI: [12.29; 26.35]; 94.7%; 27 studies; n = 3959 | 18.29 |
| 66; 95% CI: [11.46; 24.31]; I²: 87.5%; 5 studies; n = 1,096 | 16.94 |
| 45; 95% CI: [8.48; 19.53]; 89.3%; 13 studies; n = 2017 | 13.04 |
| 16/66; 95% CI: [8.52; 17.90]; I²: —; 1 study; n = 213 | 12.68 |
| 31; 95% CI: [8.96; 14.96]; I²: 37.8%; 5 studies; n = 1,075 | 11.62 |
| 16/18/45; 95% CI: [3.81; 30.05]; 93.9%; 6 studies; n = 1140 | 11.54 |
| 18/45; 95% CI: [6.27; 19.23]; I²: 89.3%; 8 studies; n = 1,433 | 11.21 |
| 18; 95% CI: [7.91; 15.10]; 83.3%; 8 studies; n = 1046 | 11 |
| 51; 95% CI: [2.96; 27.82]; I²: 87.2%; 4 studies; n = 960 | 9.78 |
| 35/53/74; 95% CI: [2.04; 25.75]; I²: —; 1 study; n = 31 | 9.68 |
| 39; 95% CI: [4.61; 17.45]; I²: 77.8%; 7 studies; n = 1,312 | 9.18 |
| 18/Other HR; 95% CI: [5.55; 13.71]; I²: 0%; 3 studies; n = 193 | 8.81 |
| 16/18; 95% CI: [4.43; 13.53]; I²: 83.0%; 3 studies; n = 720 | 7.84 |
| 33; 95% CI: [5.11; 11.75]; 68.6%; 12 studies; n = 1925 | 7.81 |
| 33/39; 95% CI: [4.35; 11.91]; I²: —; 1 study; n = 213 | 7.51 |
| 68; 95% CI: [2.99; 15.83]; I²: 69.6%; 8 studies; n = 1,006 | 7.14 |
| 59; 95% CI: [3.05; 12.58]; I²: 72.4%; 6 studies; n = 1,250 | 6.3 |
| 16/33; 95% CI: [1.84; 18.41]; I²: 84.4%; 2 studies; n = 507 | 6.13 |
| 56; 95% CI: [2.45; 13.42]; I²: 89.1%; 6 studies; n = 1,250 | 5.87 |
| 16/45; 95% CI: [1.25; 18.99]; I²: 90.0%; 4 studies; n = 613 | 5.22 |
| 33/35/52/58; 95% CI: [1.70; 13.33]; I²: 27.7%; 2 studies; n = 121 | 5.03 |
| 18/45/Other HR; 95% CI: [2.78; 8.67]; I²: 62.5%; 2 studies; n = 353 | 4.95 |
| 16/18/33; 95% CI: [2.61; 8.39]; I²: 81.8%; 8 studies; n = 1,301 | 4.71 |
| 18/33; 95% CI: [1.28; 14.99]; I²: 90.0%; 2 studies; n = 507 | 4.58 |
| 18/45/97; 95% CI: [0.56; 15.47]; I²: —; 1 study; n = 44 | 4.55 |
| 33/45; 95% CI: [0.39; 22.88]; I²: 85.2%; 2 studies; n = 507 | 3.29 |
| 53/35/74; 95% CI: [0.08; 16.70]; I²: —; 1 study; n = 31 | 3.23 |
| 45/97; 95% CI: [0.06; 12.02]; I²: —; 1 study; n = 44 | 2.27 |
| 57/82; 95% CI: [0.06; 12.02]; I²: —; 1 study; n = 44 | 2.27 |
| 52/58; 95% CI: [0.06; 12.02]; I²: —; 1 study; n = 44 | 2.27 |
| 16/31; 95% CI: [0.06; 12.02]; I²: —; 1 study; n = 44 | 2.27 |
| 33/35; 95% CI: [0.06; 12.02]; I²: —; 1 study; n = 44 | 2.27 |
| 13/52; 95% CI: [0.06; 12.02]; I²: —; 1 study; n = 44 | 2.27 |
| 18/68; 95% CI: [0.06; 12.02]; I²: —; 1 study; n = 44 | 2.27 |
| 33/54; 95% CI: [0.06; 12.02]; I²: —; 1 study; n = 44 | 2.27 |
| 16/18/Other HR; 95% CI: [0.84; 5.77]; I²: 0%; 2 studies; n = 180 | 2.22 |
| 16/33/45; 95% CI: [0.02; 4.00]; I²: —; 1 study; n = 137 | 0.73 |
| 18/33/45; 95% CI: [0.02; 4.00]; I²: —; 1 study; n = 137 | 0.73 |

**Supplementary Fig. 1** Trend of HPV strains identified among human papillomavirus-positive women in Cameroon

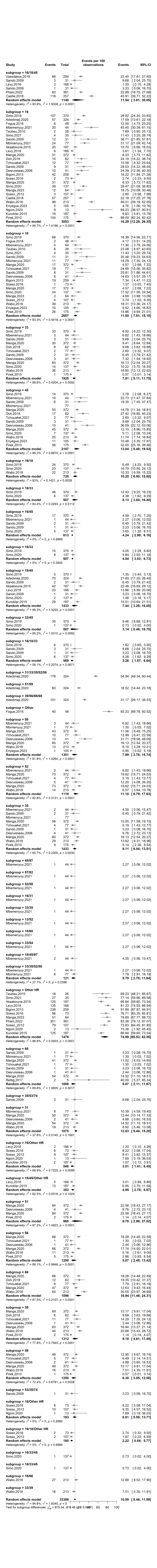

**Continue…**

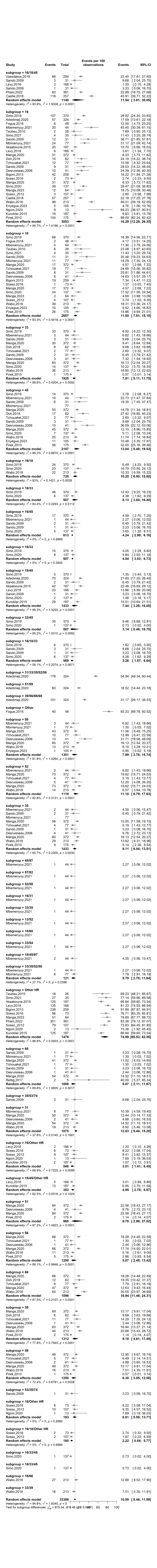

**Continue…**

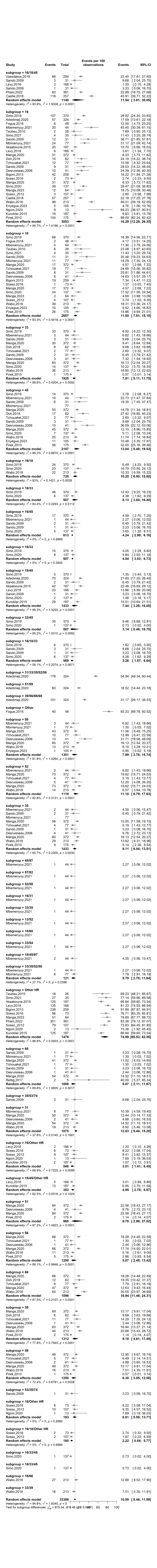

**Continue…**

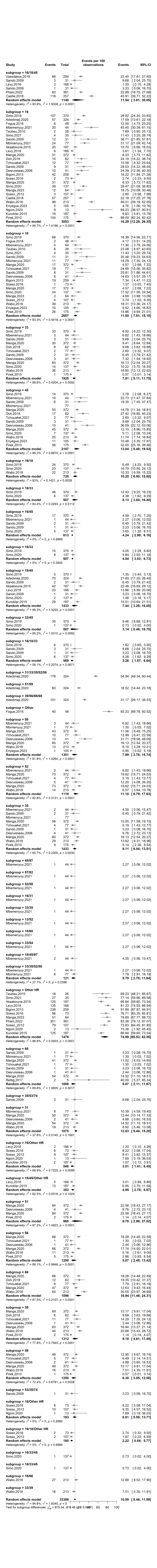

**Supplementary Fig. 2** Genotypic profile of human papillomavirus strains identified among HPV-positive women in Cameroon

**Histological profile**

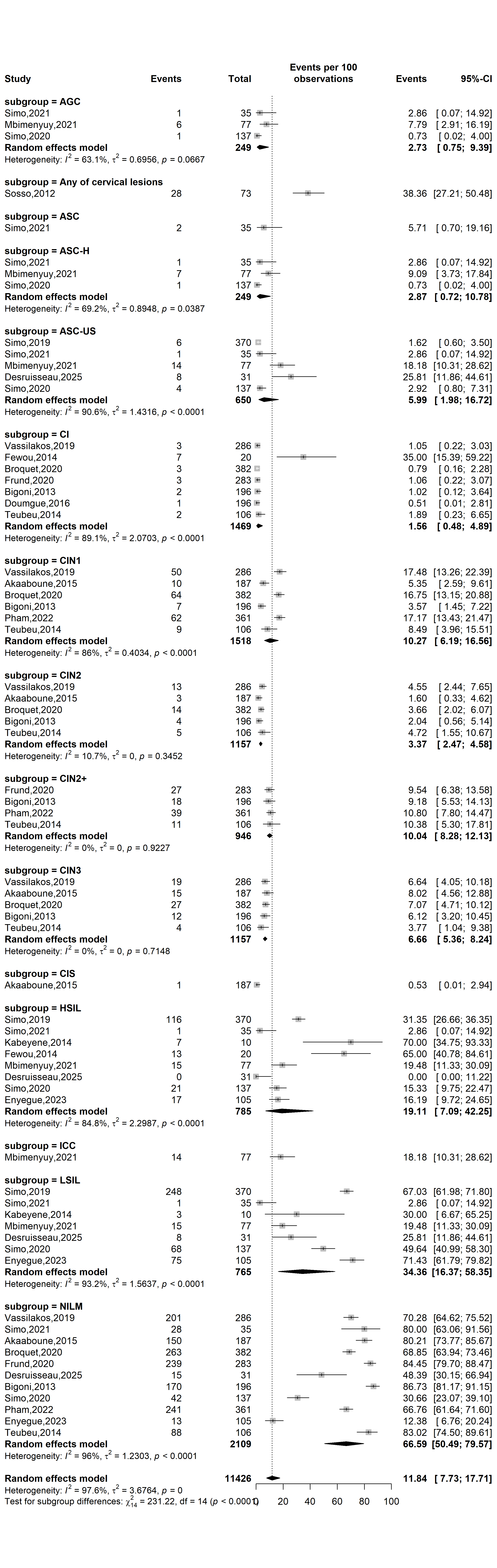

**Continue…**

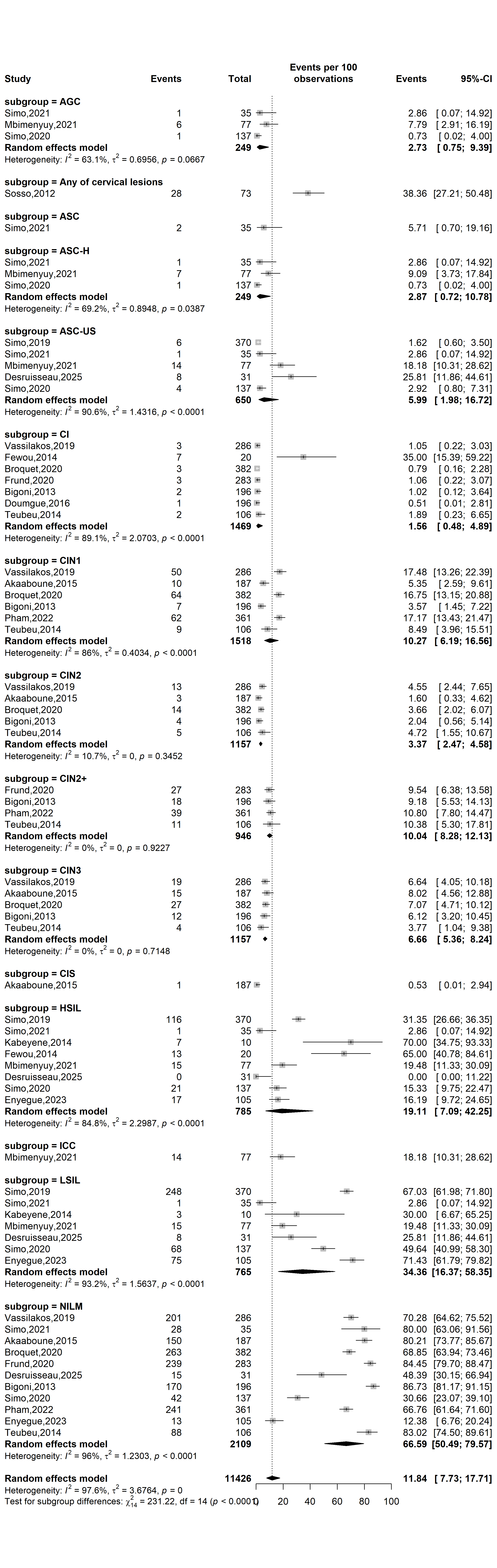

**Supplementary Fig. 3** Histological patterns of the uterine cervix among HPV positive women in Cameroon

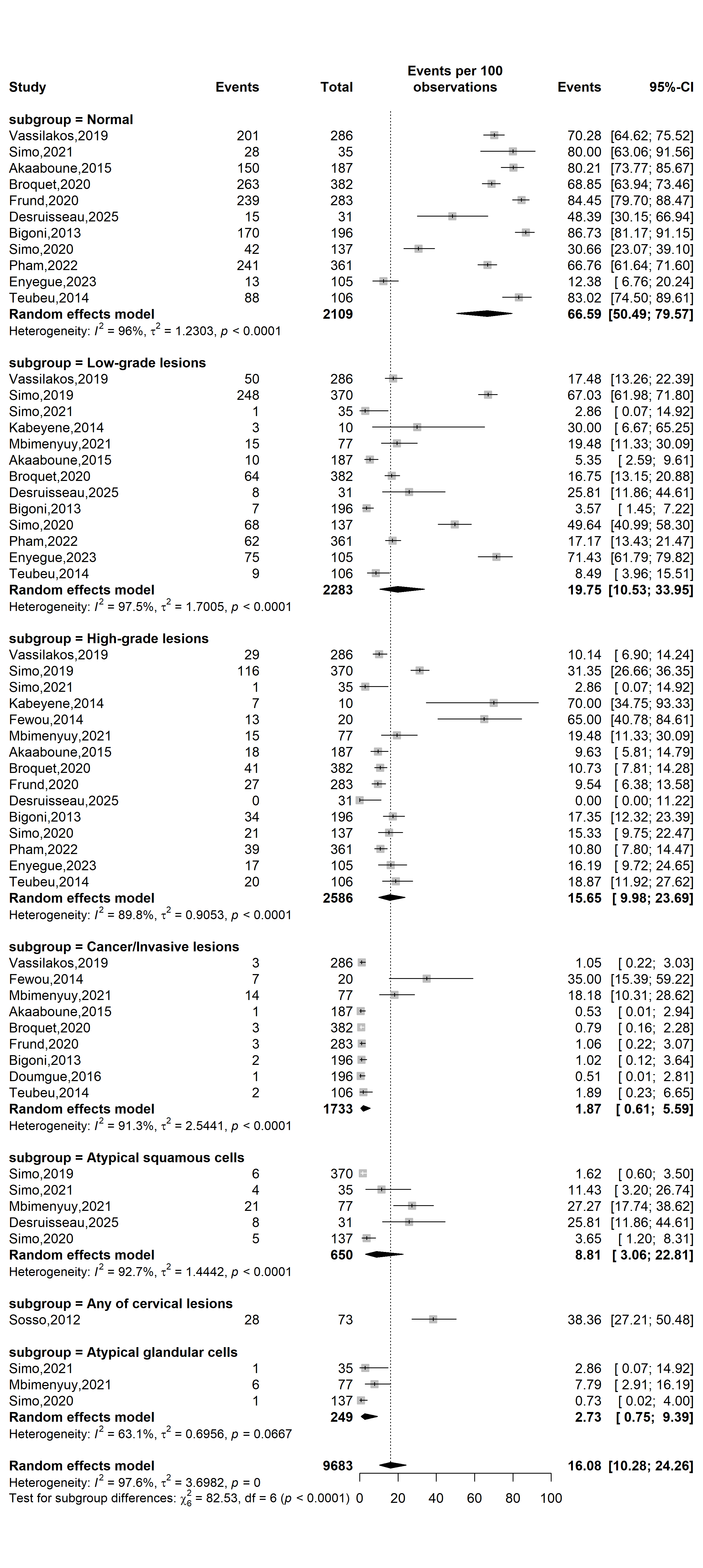

**Supplementary Fig. 4** Histological patterns of the uterine cervix among HPV positive women in Cameroon
