## Supplementary material for "Prevalence of high-risk human papillomavirus genotypes and associated HIV co-infection in Cameroon: a systematic review and meta-analysis": Addition file 1

**Supplementary Table 1** Searching strategy by database

| **Database** | **Search string** | **Number of entries** |
| --- | --- | --- |
| **Pubmed** | ("human papillomavirus"[tiab] OR HPV[tiab] OR "cervical cancer"[tiab] OR "uterine cervical neoplasms"[Mesh]) AND (Cameroon[tiab] OR "Cameroon"[Mesh]) | 121 |
| **Scopus** | TITLE-ABS-KEY (("human papillomavirus" OR HPV OR "uterine cervical neoplasm") AND (Cameroon OR Cameroonian)) | 120 |
| **Web of sciences** | TS= ("human papillomavirus" OR HPV OR "cervical cancer" OR OR "uterine cervical cancer") AND TS= (Cameroon OR Cameroonian) | 217 |
| **Embase** | ('human papillomavirus':ti,ab,kw OR 'HPV':ti,ab,kw OR 'cervical cancer':ti,ab,kw OR 'uterine cervical neoplasms':ti,ab,kw) AND ('Cameroon':ti,ab,kw OR 'Cameroonian':ti,ab,kw) | 196 |
| **Cochrane Library** | (human papillomavirus OR HPV OR cervical cancer) AND (Cameroon OR Cameroun) | 37 |
| **AJOL** | (human papillomavirus OR HPV OR cervical cancer) AND (Cameroon OR Cameroun) | 33 |
| **Health Sciences and Disease** | (human papillomavirus OR HPV OR cervical cancer) AND (Cameroon OR Cameroun) | 9 |

**Subgroup analysis of high-risk HPV infection prevalence**

**Study period**

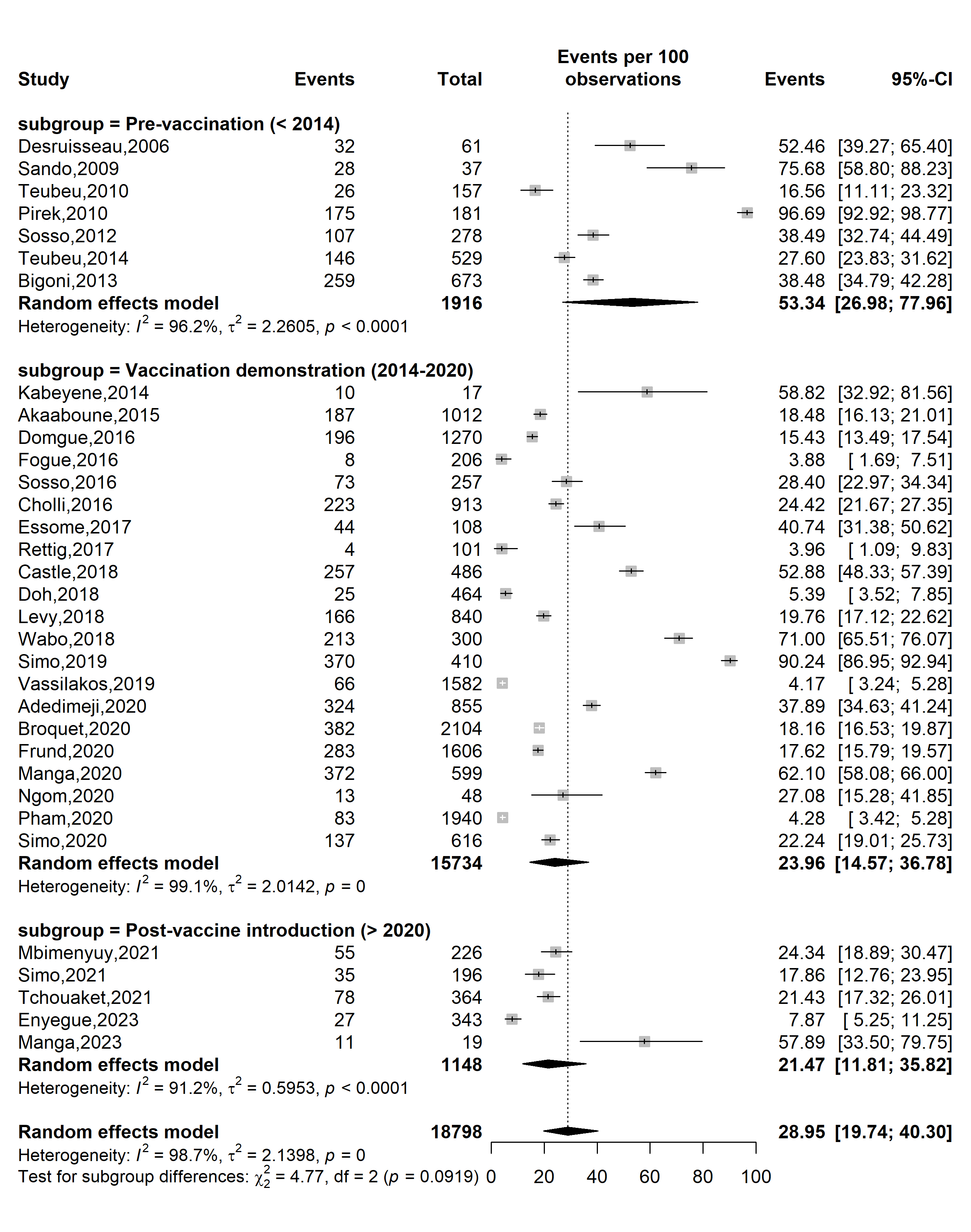

**Supplementary Fig. 1** Pooled high-risk HPV prevalence according to specific HPV vaccine introduction timeframe in Cameroon

**Study design**

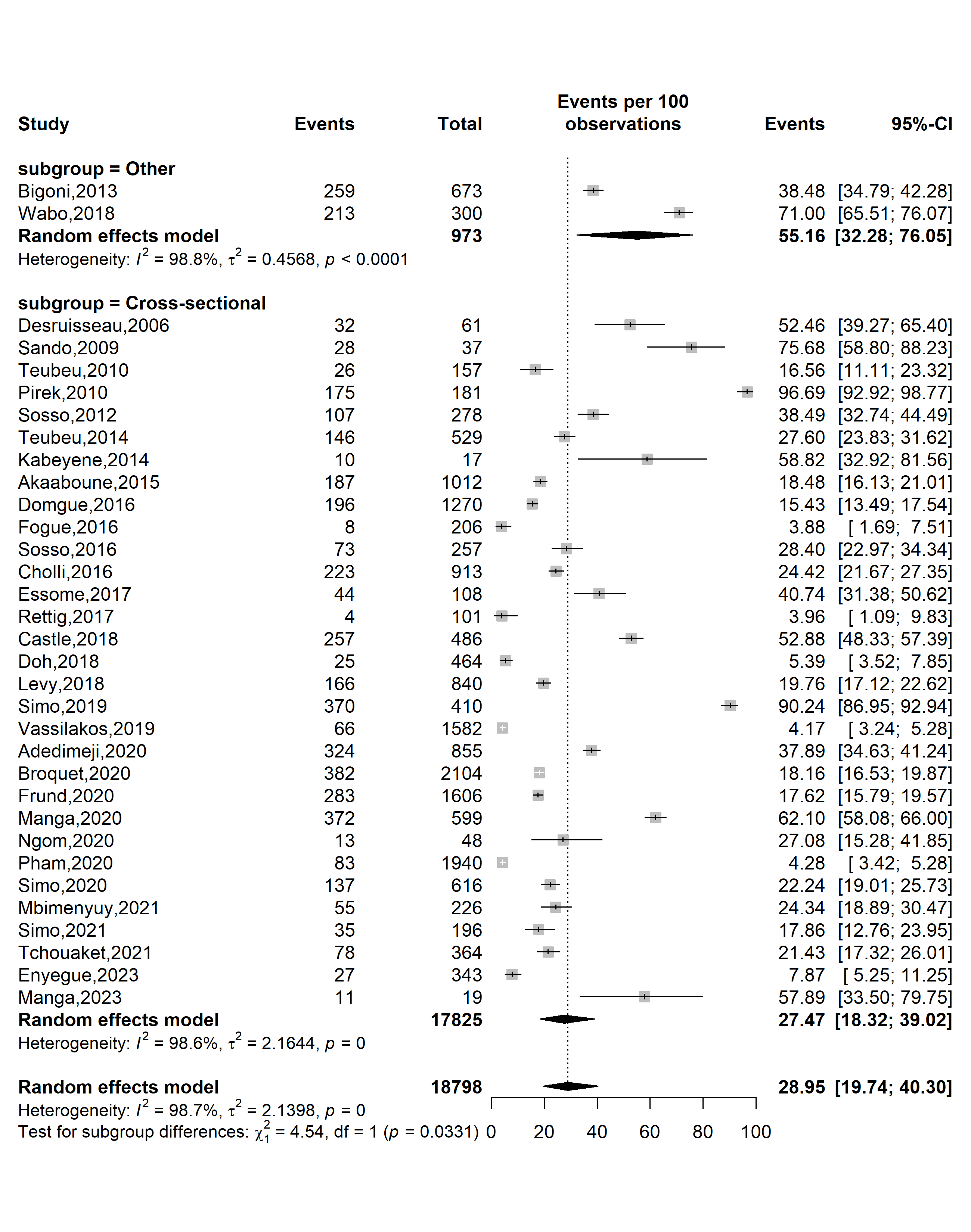

**Supplementary Fig. 2** Pooled high-risk HPV prevalence in Cameroon by study design

**Study setting**

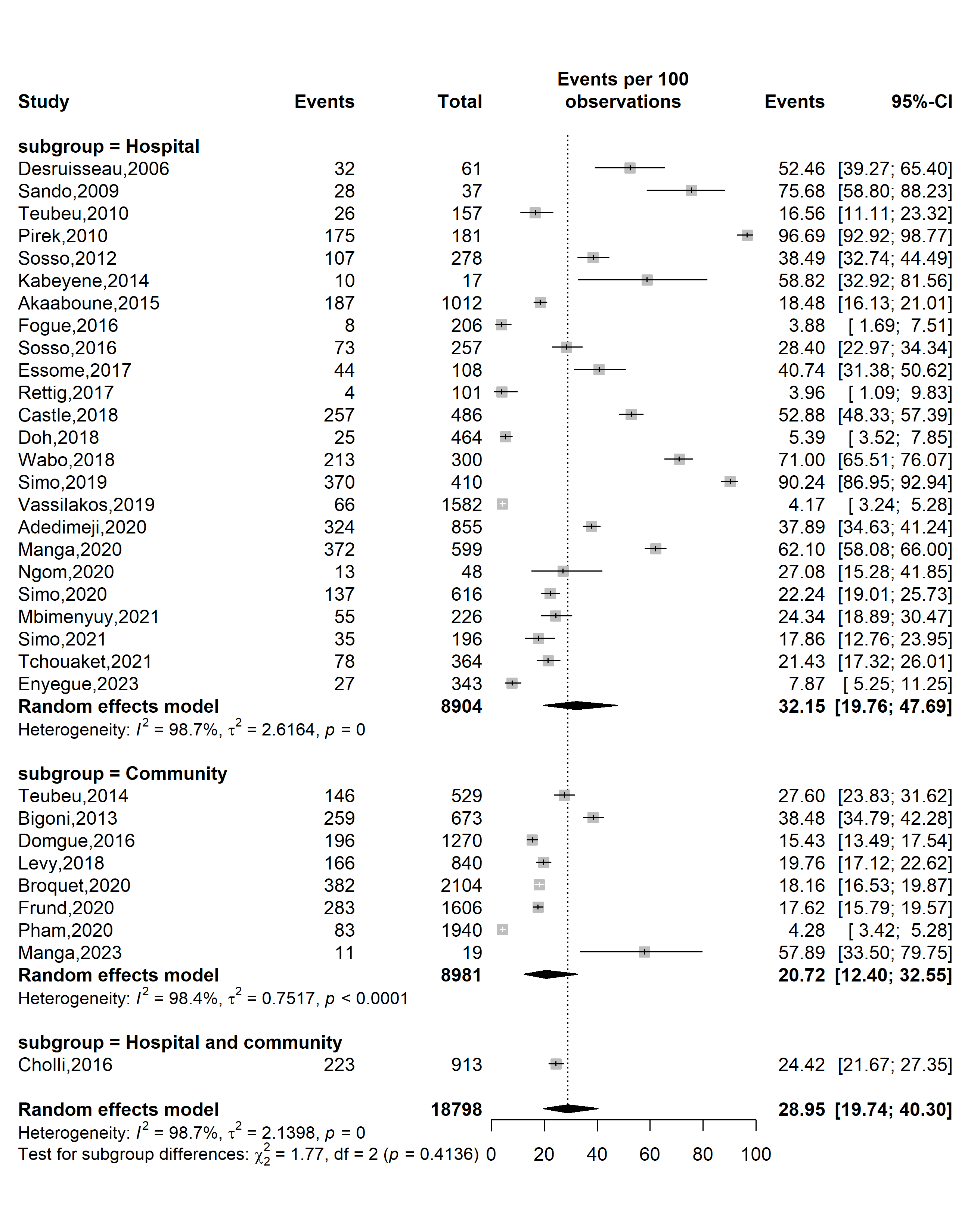

**Supplementary Fig. 3** Pooled high-risk HPV prevalence in Cameroon by study setting

**Study site**

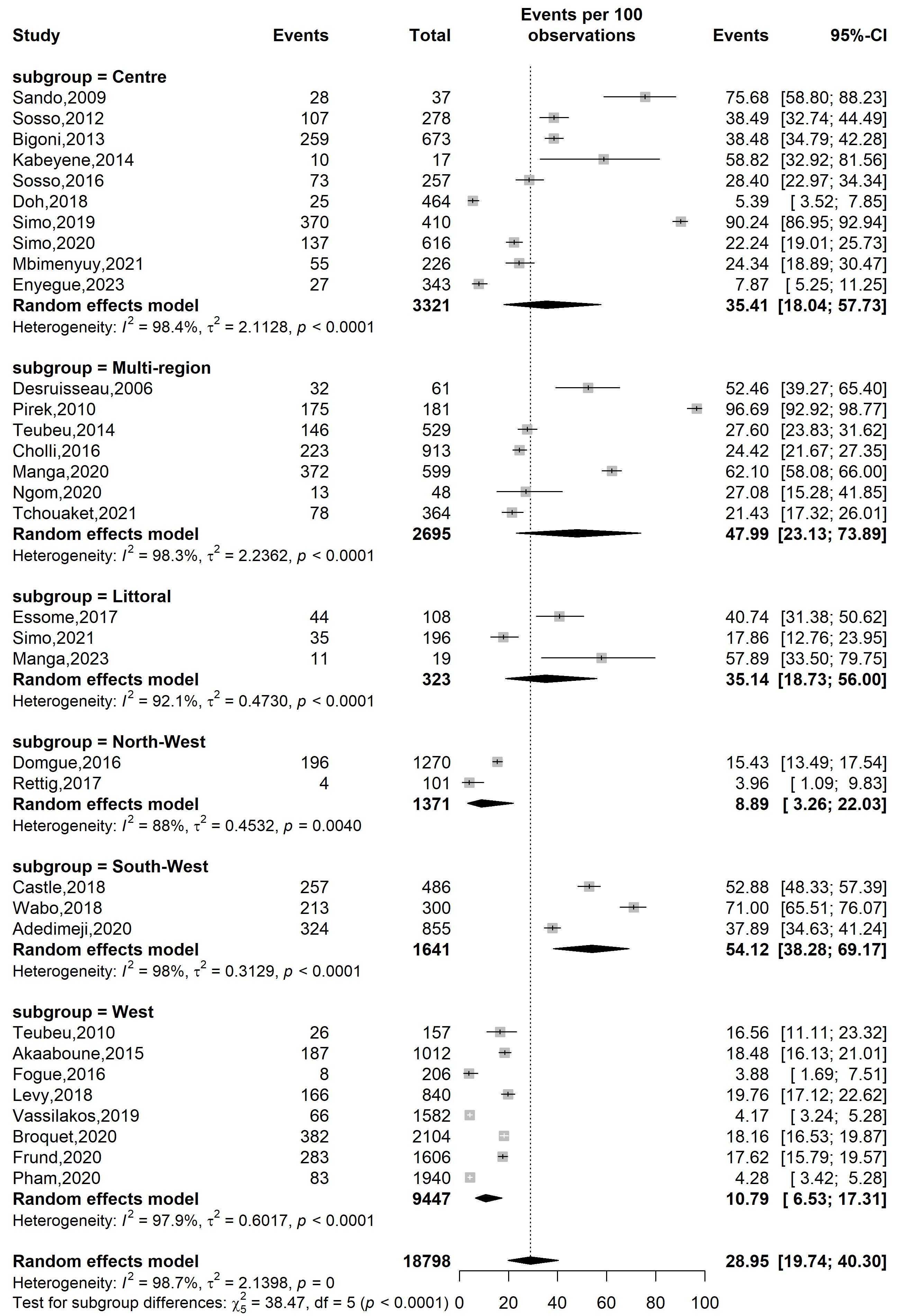

**Supplementary Fig. 4** Pooled high-risk HPV prevalence in Cameroon by study site 1

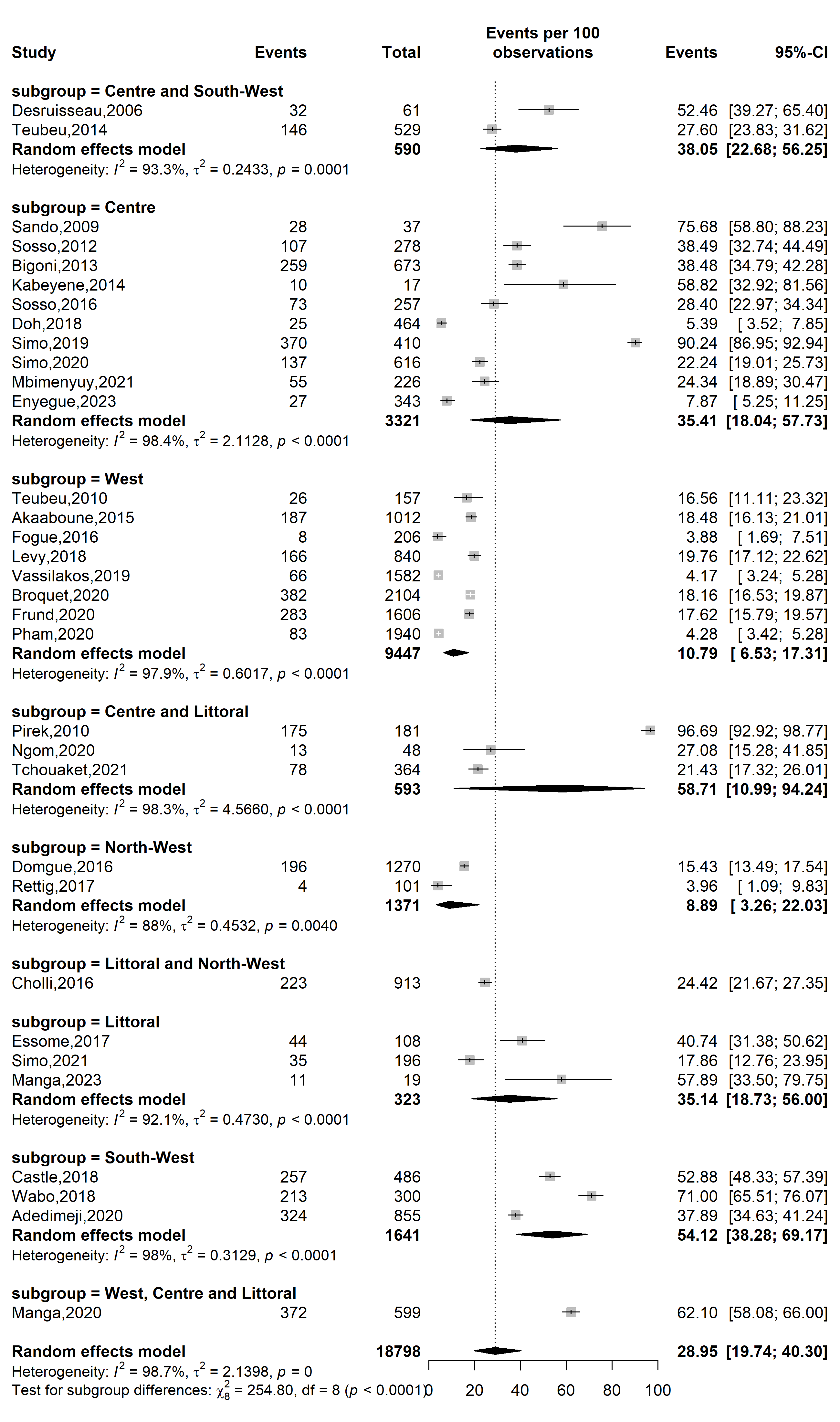

**Supplementary Fig. 5** Pooled high-risk HPV prevalence in Cameroon by study site 2

**Sampling method**

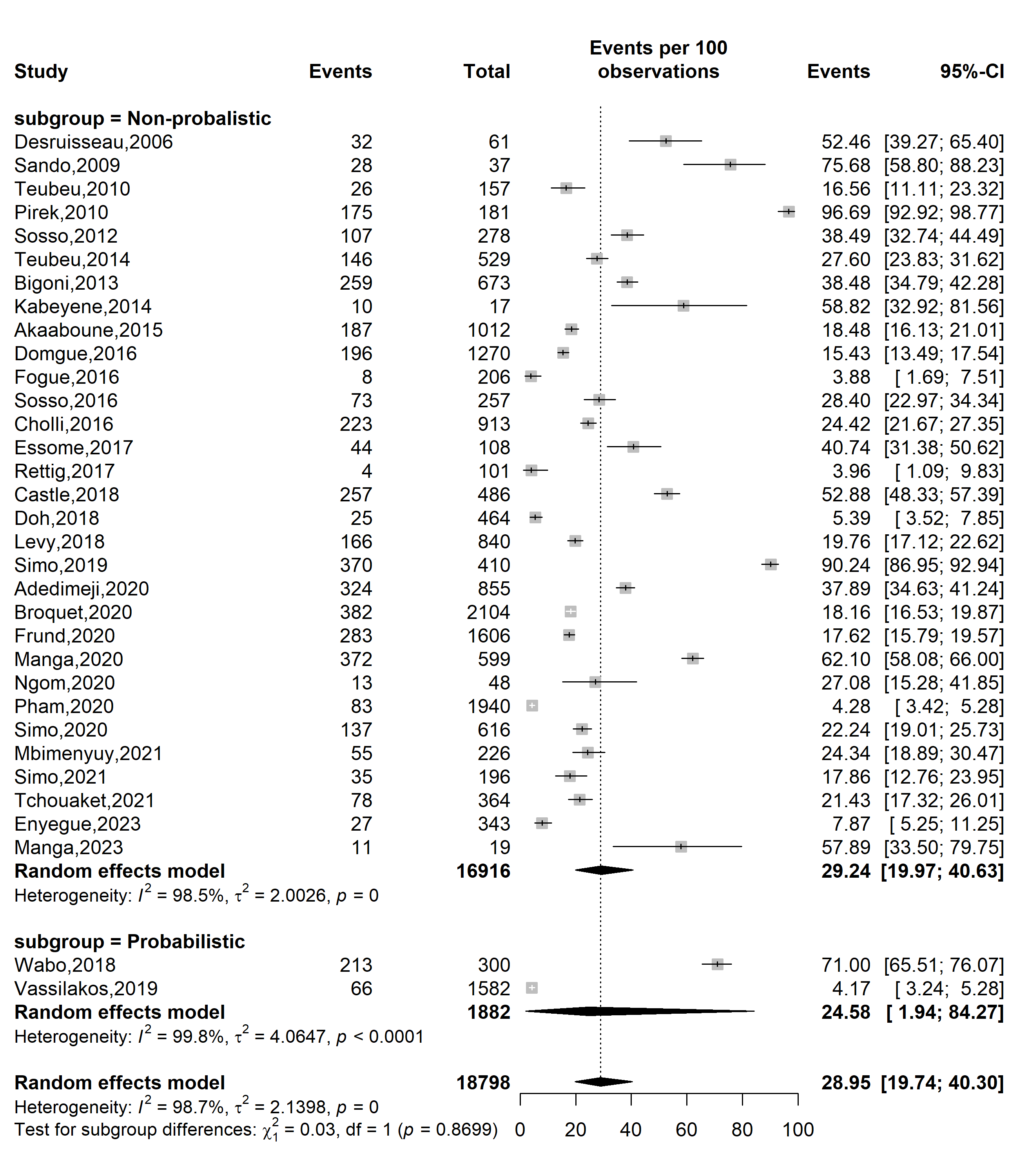

**Supplementary Fig. 6** Pooled high-risk HPV prevalence in Cameroon by sampling method

**Participant**

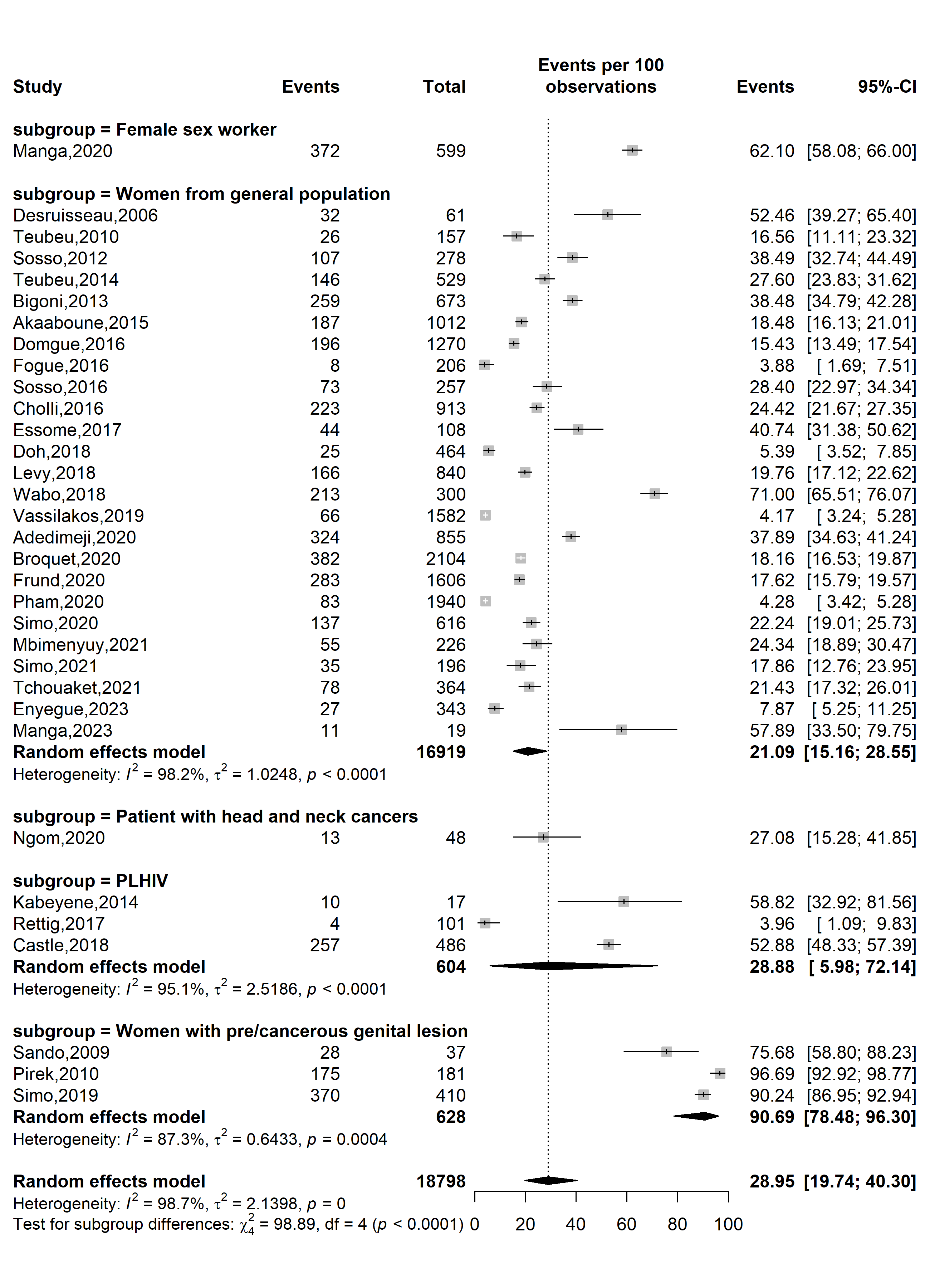

**Supplementary Fig. 7** Pooled high-risk HPV prevalence in Cameroon by type of participants

**Sensitivity analysis**

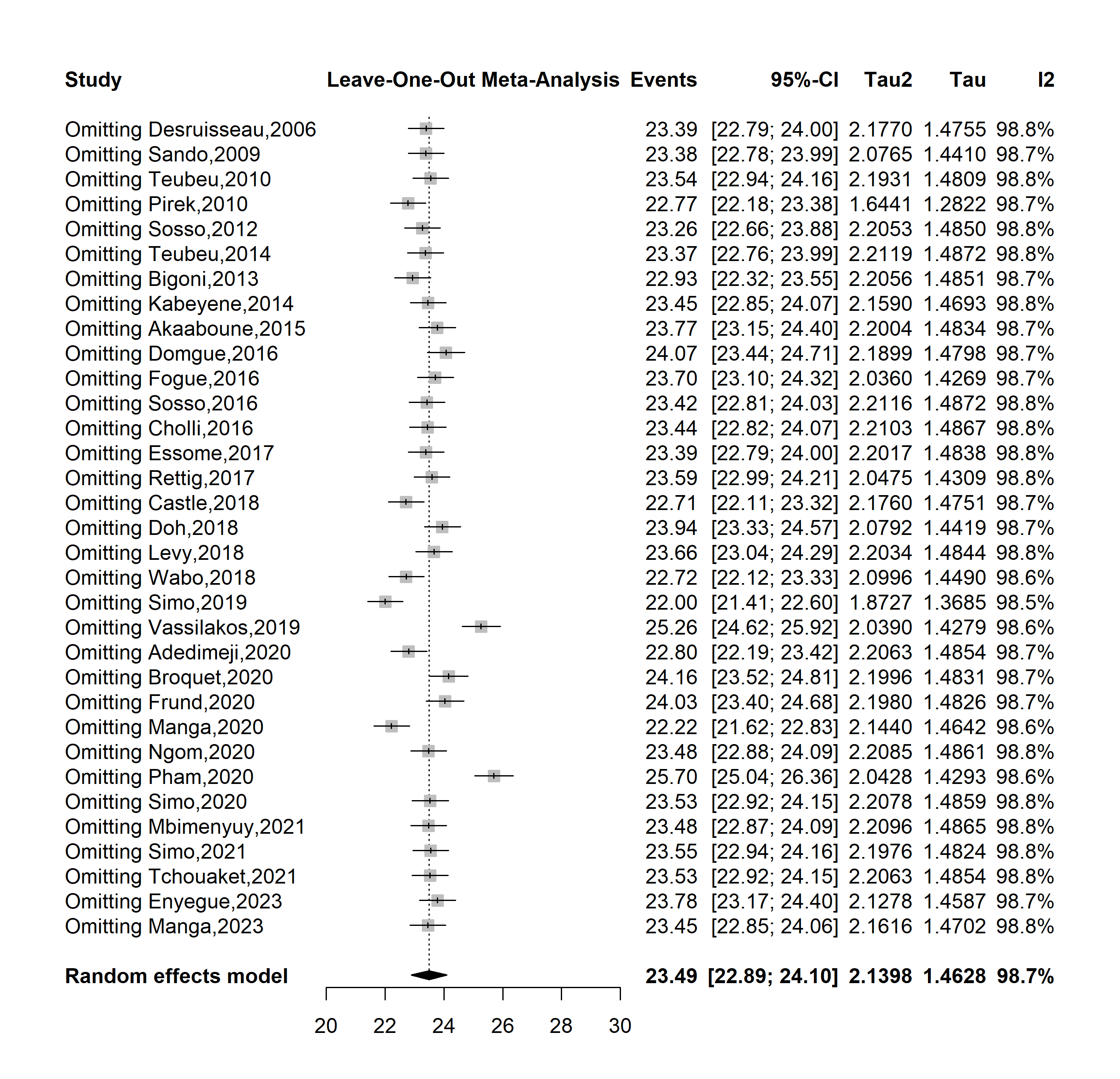

**Supplementary Fig. 8** Sensitivity analysis of high-risk HPV prevalence in Cameroon

**Publication bias assessment**

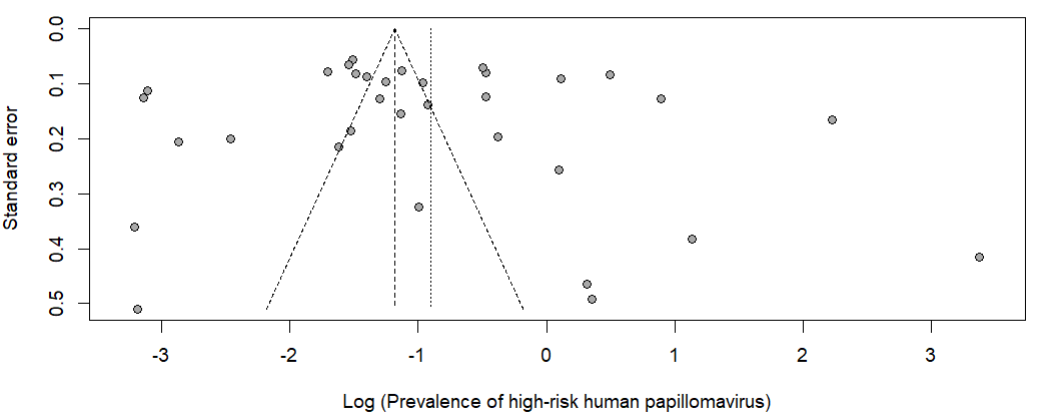

Egger’s test *p*-value = 0.570

Begg’s test *p*-value = 0.495

**Supplementary Fig. 9** Funnel plot assessing publication bias among studies assessing the high-risk prevalence in Cameroon

**Subgroup analysis of HIV infection prevalence among HPV-positive cases**

**Study year**

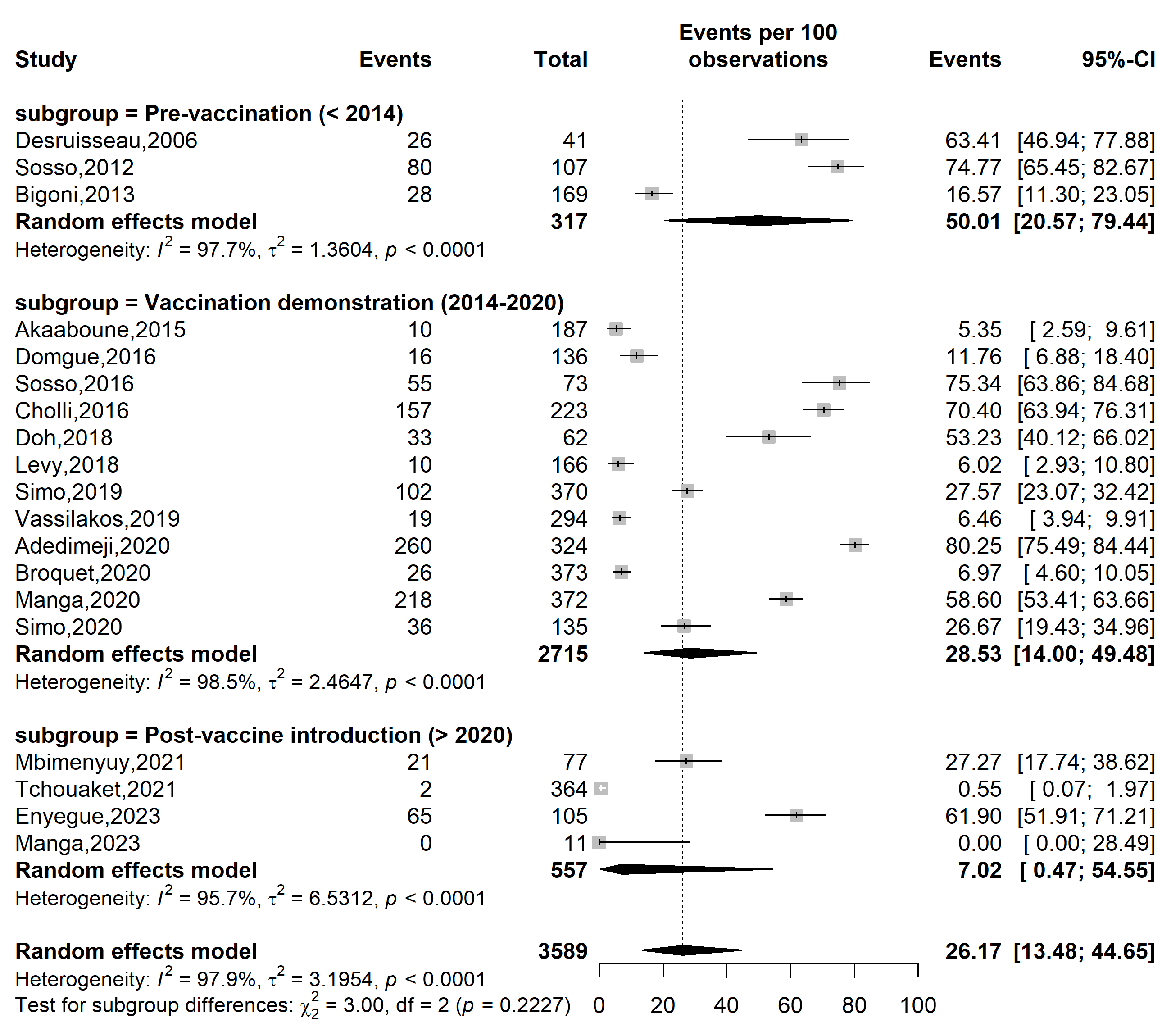

**Supplementary Fig. 10** Pooled HIV infections among HPV-positive cases according to vaccine introduction timeframe in Cameroon

**Study design**

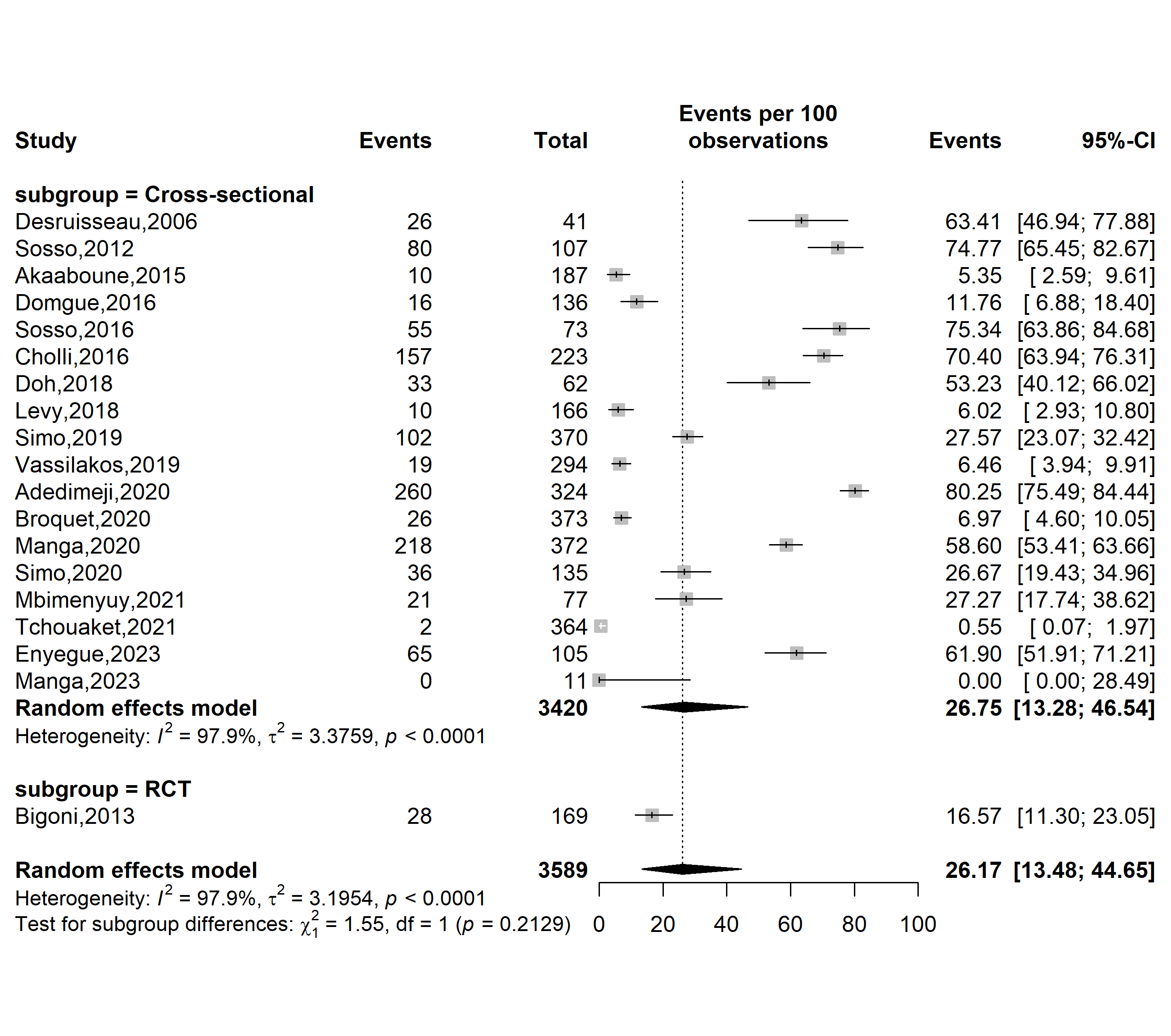

**Supplementary Fig. 11** Pooled HIV infections among HPV-positive cases in Cameroon by study designs (RCT: randomized control trial)

**Study setting**

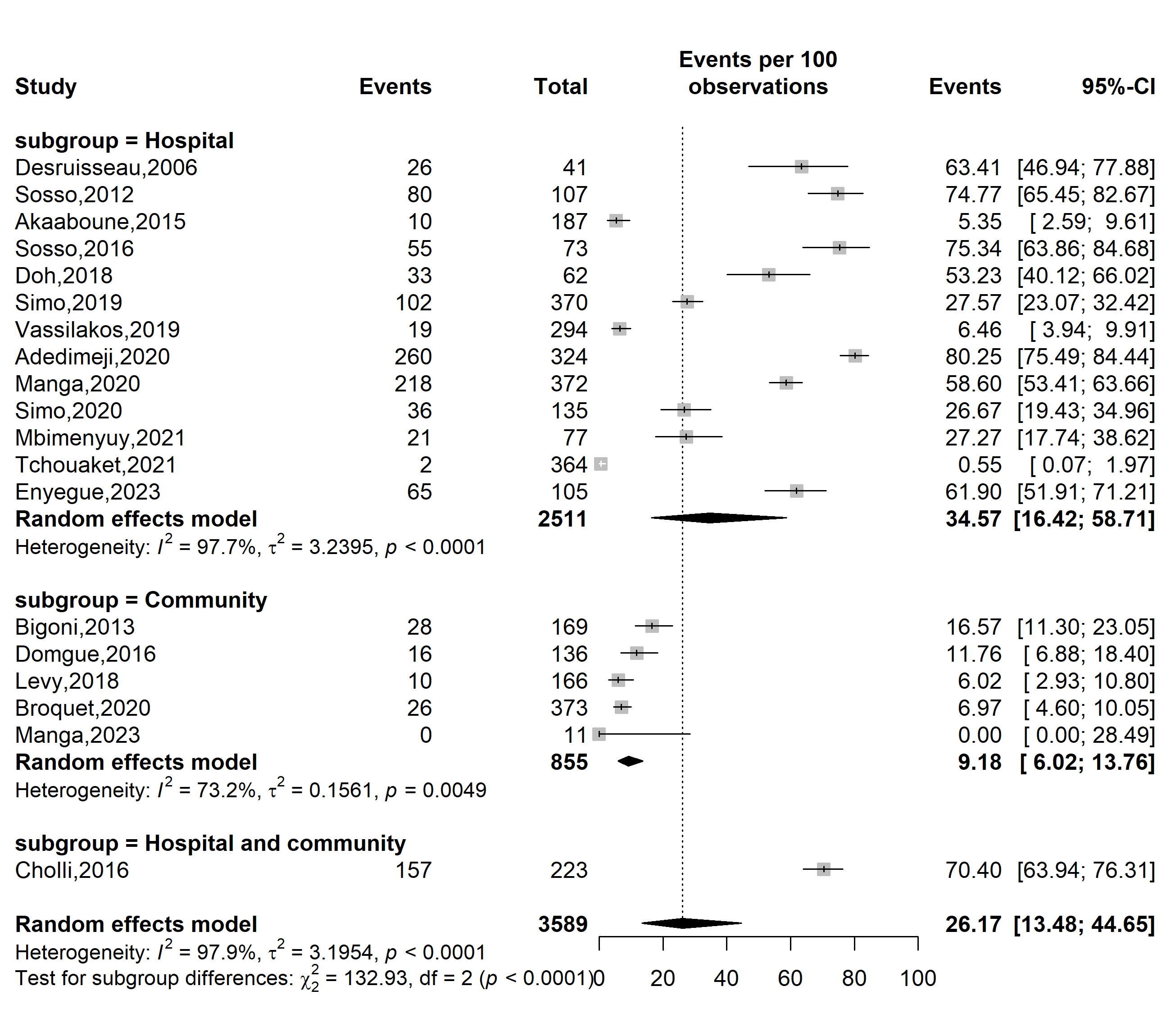

**Supplementary Fig. 12** Pooled HIV infections among HPV-positive cases in Cameroon by study settings

**Study site**

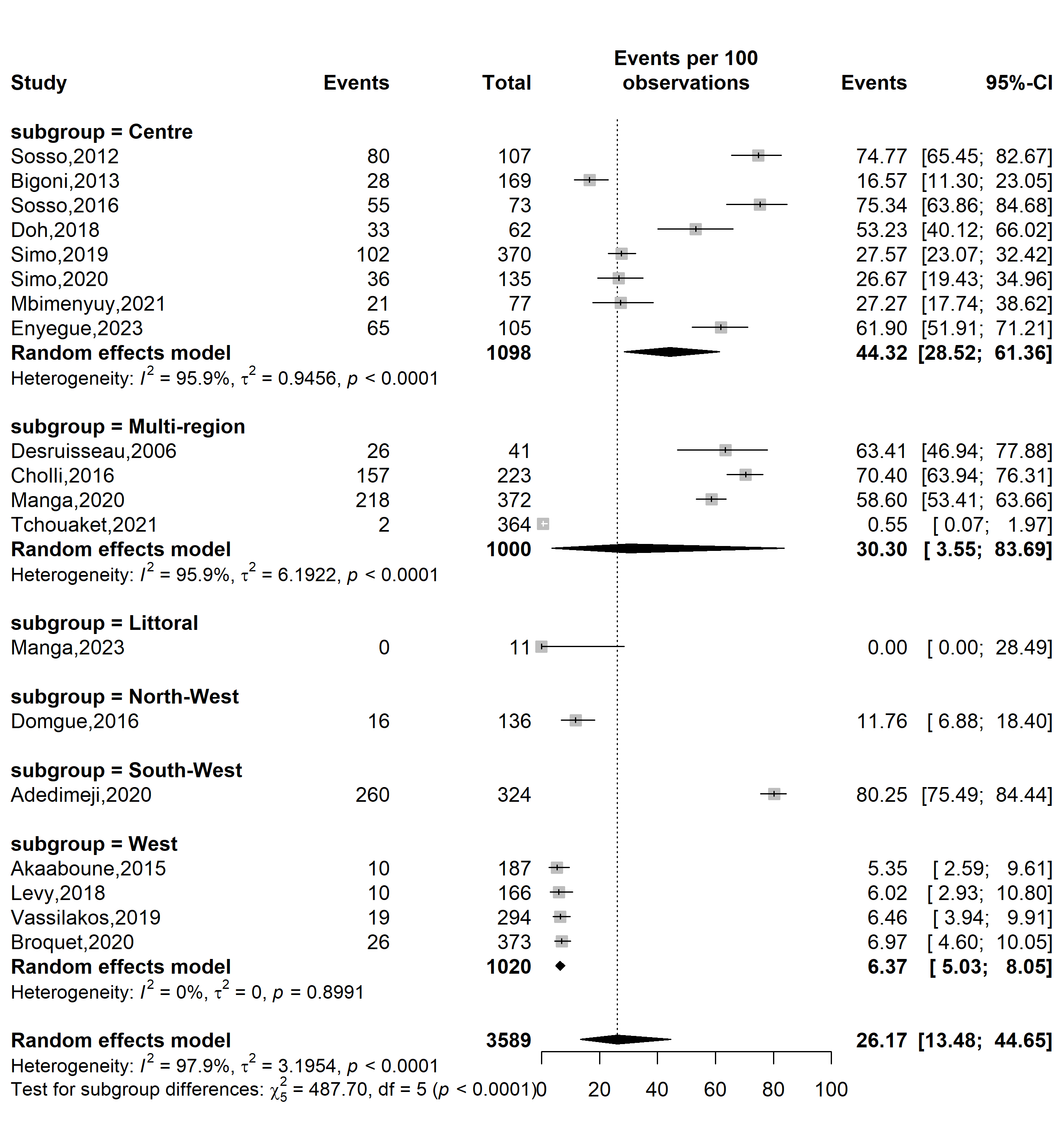

**Supplementary Fig. 13** Pooled HIV infections among HPV-positive cases in Cameroon by study site 1

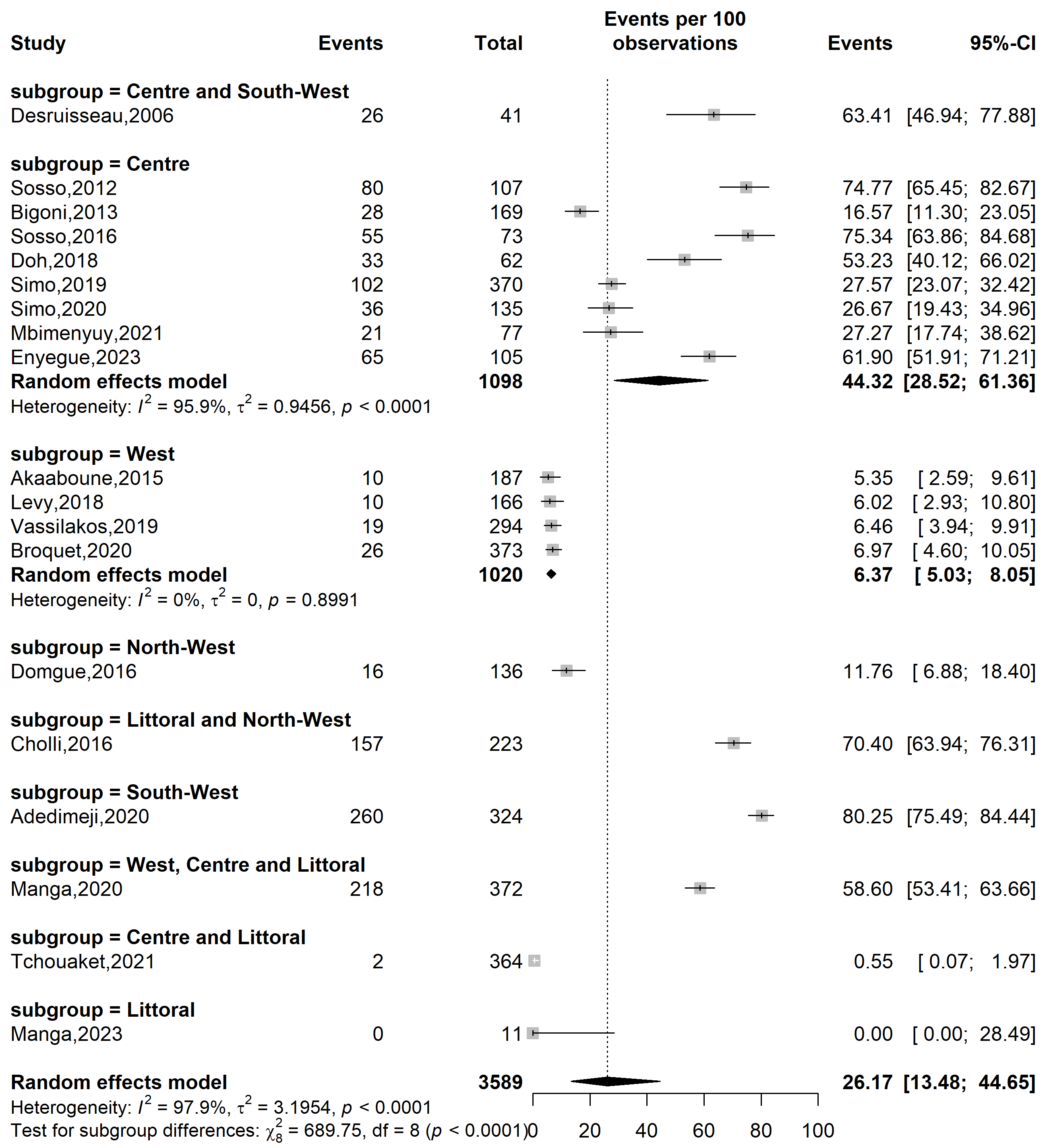

**Supplementary Fig. 14** Pooled HIV infections among HPV-positive cases in Cameroon by study site 2

**Sampling method**

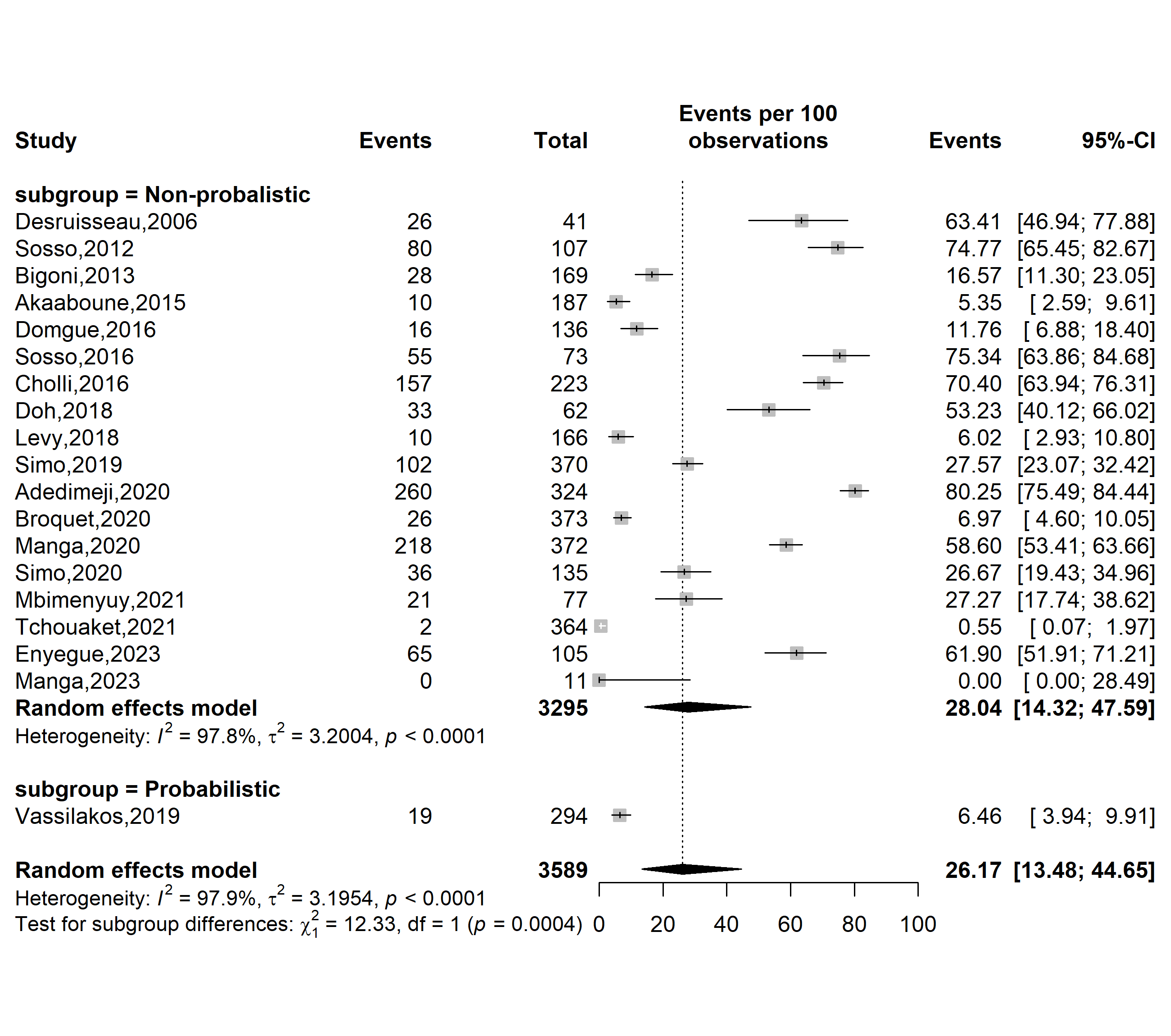

**Supplementary Fig. 15** Pooled HIV infections among HPV-positive cases in Cameroon by sampling method

**Participants**

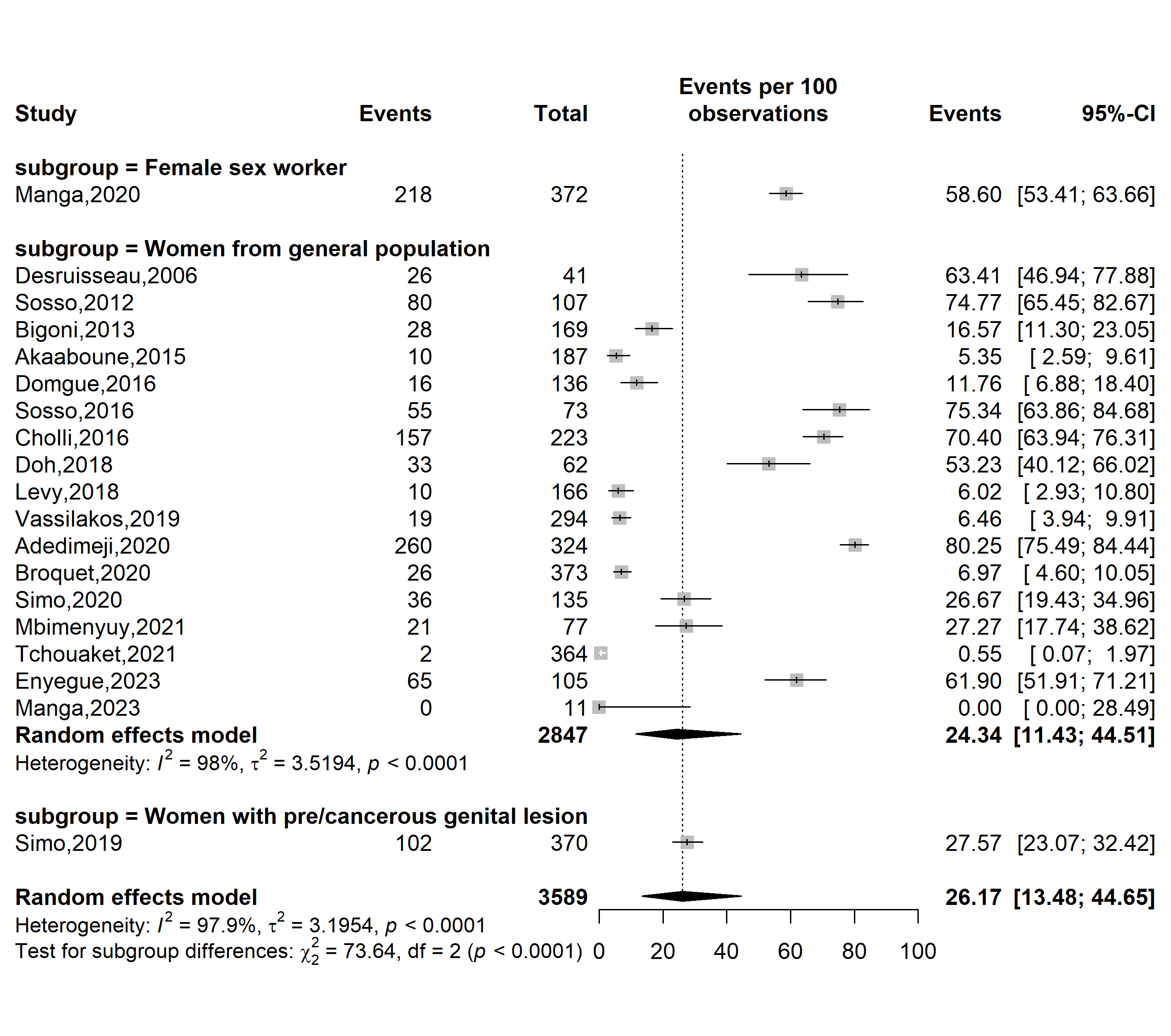

**Supplementary Fig. 16** Pooled HIV infections among HPV-positive cases in Cameroon by study participants

**Sensitivity analysis**

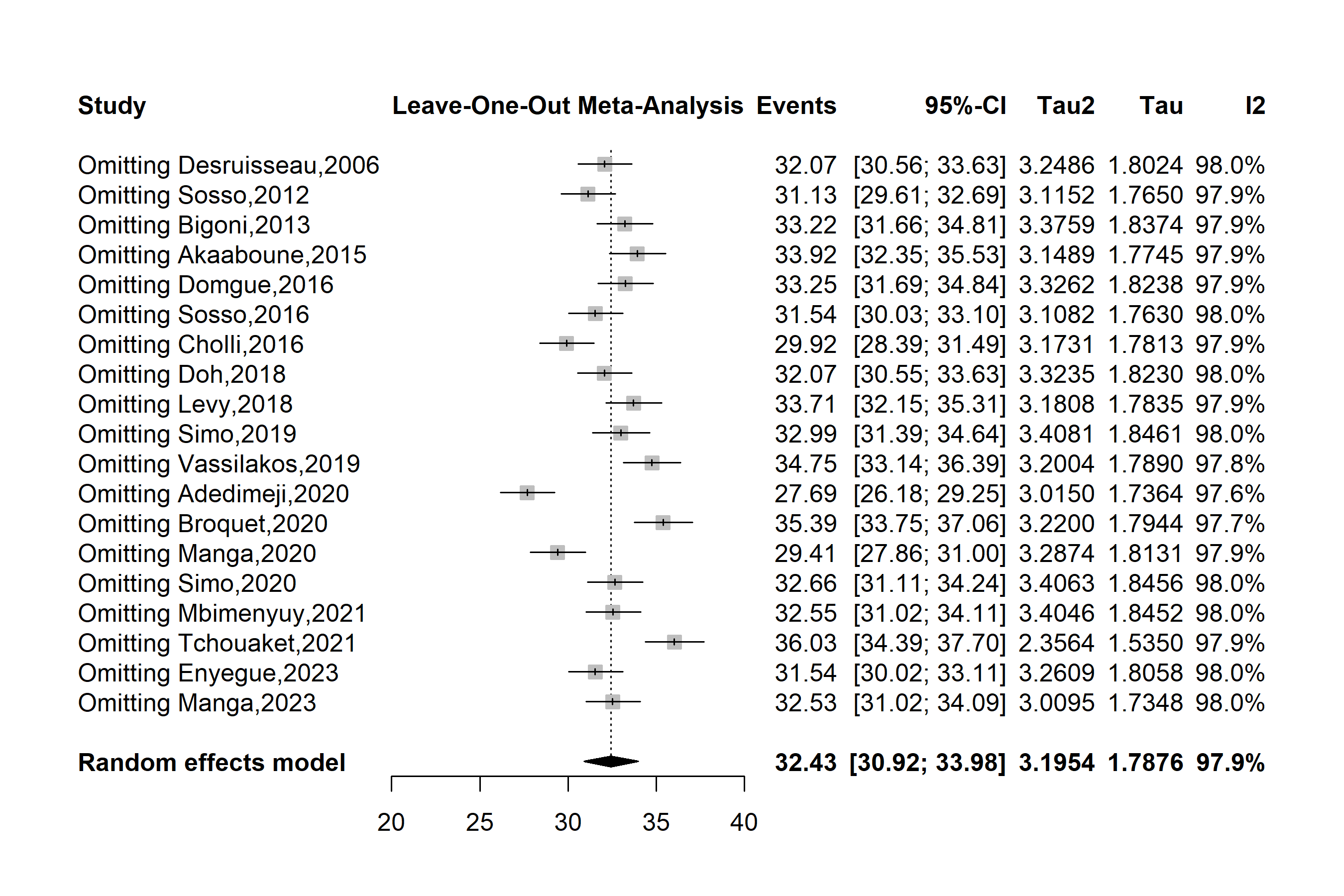

**Supplementary Fig. 17** Sensitivity analysis of the pooled HIV infections among HPV-positive cases in Cameroon

**Publication bias assessment**

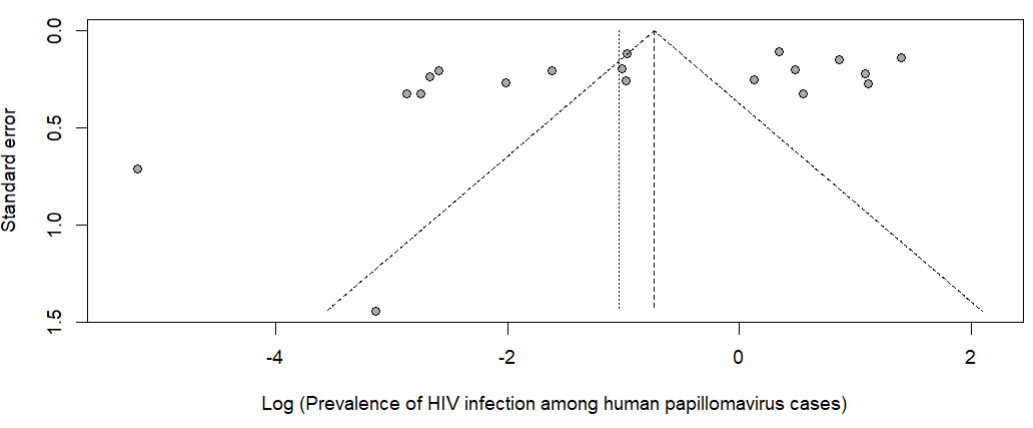

Egger’s test *p*-value = 0.071

Begg’s test *p*-value = 0.100

**Supplementary Fig. 18** Funnel plot assessing publication bias among studies assessing the HIV prevalence among HPV-positive individuals in Cameroon

**Sensitivity analysis**

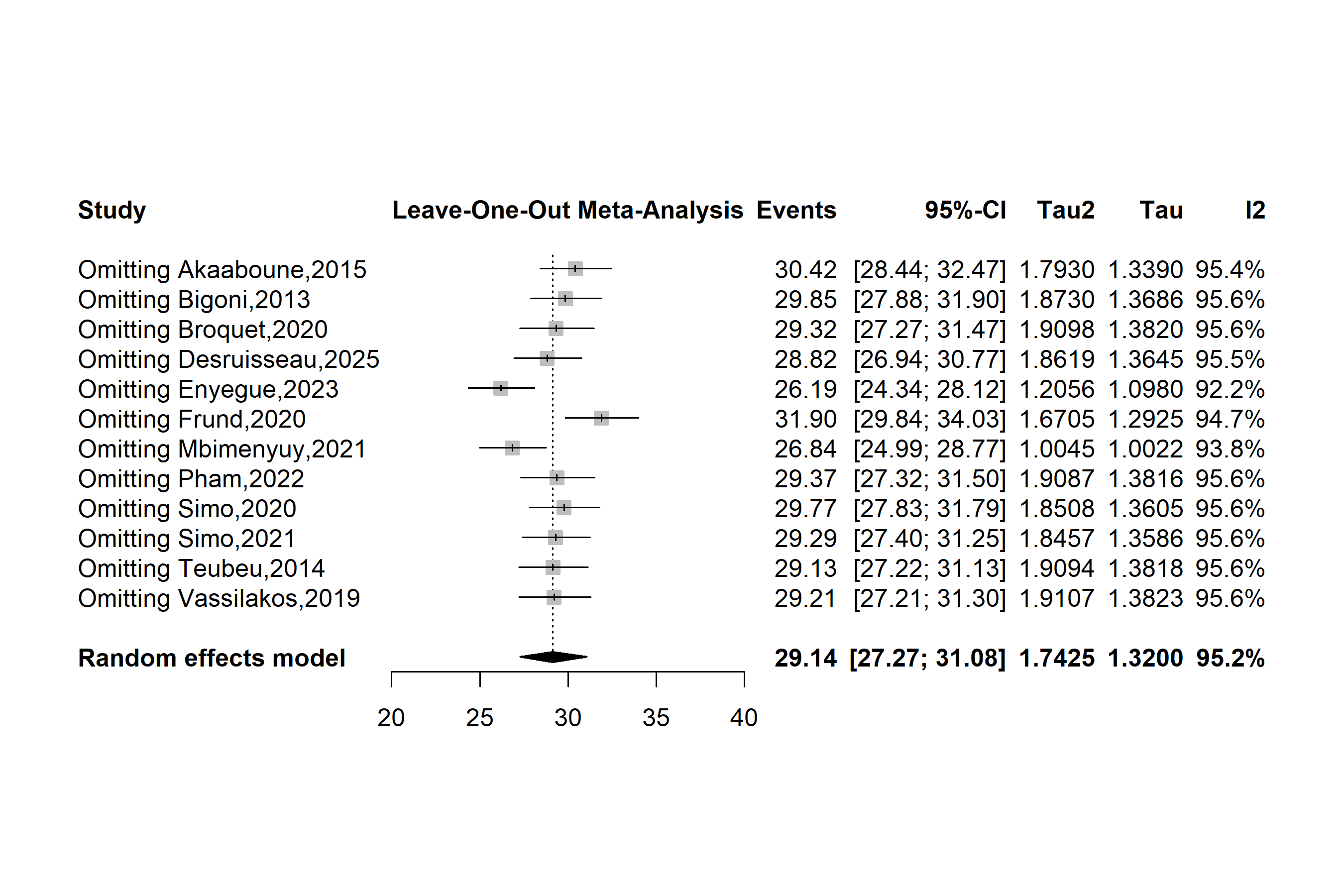

**Supplementary Fig. 19** Sensitivity analysis of the pooled HIV infections among HPV-positive cases in Cameroon

**Publication bias assessment**

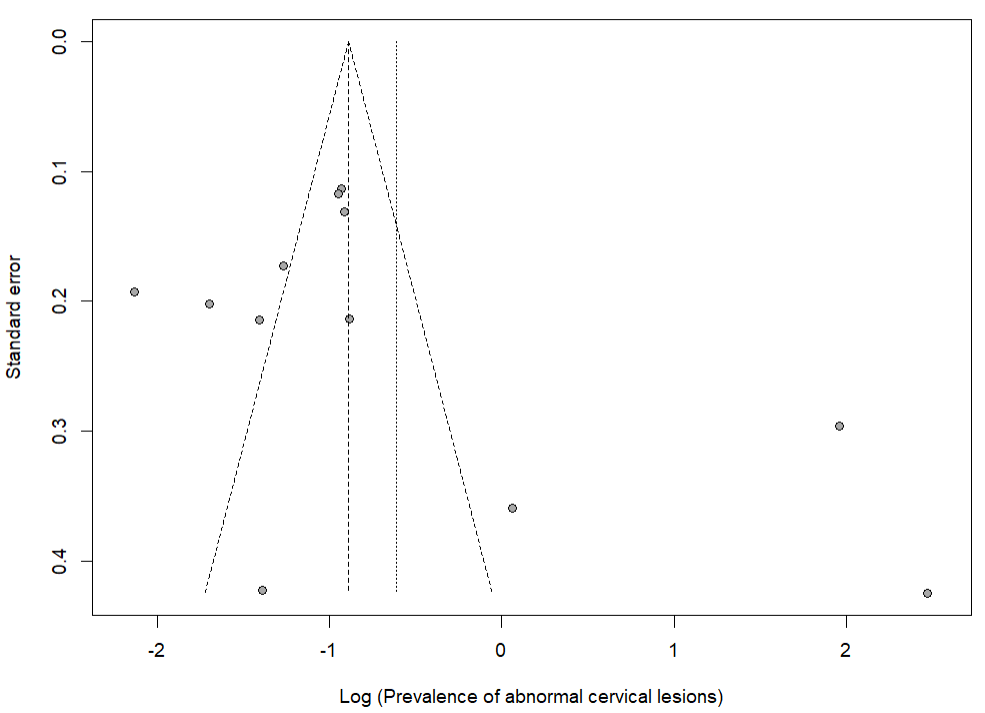

Egger’s test *p*-value = 0.214

Begg’s test *p*-value = 0.337

**Supplementary Fig. 18** Funnel plot assessing publication bias among studies assessing the prevalence of abnormal cervical lesions among HPV-positive women in Cameroon
